# Interrogation of T Cell-Enriched Tumors Reveals Prognostic and Immunotherapeutic Implications of Polyamine Metabolism

**DOI:** 10.1101/2021.10.18.21265179

**Authors:** R. Alex Harbison, Rajeev Pandey, Michael Considine, Robert D. Leone, Tracy Murray-Stewart, Rossin Erbe, Raj Mandal, Mark Burns, Robert Casero, Tanguy Seiwert, Carole Fakhry, Drew Pardoll, Elana Fertig, Jonathan D. Powell

**Author notes:** **Correspondence to:** Jonathan D. Powell, MD, PhD, Calico Life Sciences LLC, 1170 Veterans Blvd., South San Francisco, CA, 94080. Elana Fertig, PhD, Suite 1101E, 550 N. Broadway, Baltimore, MD, 21205.

## Abstract

**Background:** The presence of cytotoxic tumor infiltrating lymphocytes (TILs) and antigen (e.g., viral, tumor neoantigens) enhances anti-tumor immunity. However, features including recruitment of tolerogenic cell types, nutrient-depletion, and the establishment of an acidic and hypoxic microenvironment diminish anti-tumor lymphocyte function. We sought to understand why the anti-tumor immune response fails despite a favorable immune profile.

**Methods:** We leveraged human papillomavirus-related (HPV+) head and neck squamous cell carcinomas (HNSC) to address this question given their high degree of CD8^+^ T cell-infiltration and virus-derived tumor-associated antigens. We evaluated expression of 2,520 metabolic genes between HPV+ HNSCs of different prognostic phenotypes. We further tested tumor-intrinsic and -extrinsic sources of polyamine (PA) gene expression based on observations from the prior analysis. We used bulk RNAseq from The Cancer Genome Atlas (TCGA; 10 different cancers) and single cell (sc) RNAseq data from two atlases to parse immune cell contributions to polyamine gene expression. We used TCGA data and an immunotherapy-treated melanoma cohort to examine survival outcomes as a function of polyamine gene set expression.

**Results:** PA metabolism genes were upregulated in aggressive phenotype, T cell-enriched (Thi), HPV+ HNSCs. PA synthesis and transporter gene enrichment was associated with T cell infiltration, recurrent or persistent cancer, overall survival status, primary site, molecular subtype, and *MYC* genomic alterations. PA synthesis and transport gene sets were more highly expressed in HPV-compared to HPV+ HNSCs. Bulk and scRNAseq data from HPV+ HNSCs demonstrated greater PA catabolism gene set expression among myeloid cells. A combined PA gene set comprised of genes involved in PA synthesis and transport was negatively correlated with cytotoxic T cell functional score across TCGA tumor types. Combined PA gene set expression was associated with greater mortality risk across five tumor types and worse survival in T cell-infiltrated, anti-PD-1-treated melanomas.

**Conclusions:** A genomic approach leveraging T cell-infiltrated, immunogenic HPV+ HNSCs revealed an association between polyamine metabolism, anti-tumor immunity, and prognosis across several cancer types. These data address hurdles to anti-tumor immunity and immunotherapy and warrant further investigation of polyamines as a biomarker for targeted therapy in the context of a T cell-infiltrated microenvironment.

## INTRODUCTION

Features promoting anti-tumor immunity include the presence of cytotoxic tumor infiltrating lymphocytes (TILs) and antigen (e.g., viral, tumor neoantigens).^1-4^ However, not all immune cell infiltrated, immunogenic tumors exhibit favorable anti-tumor immunity. The tumor microenvironment (TME) diminishes anti-tumor T cell function through recruitment of tolerogenic cell types, nutrient-depletion, and the creation of an acidic and hypoxic microenvironment.^5^ Immune modulating therapies may be affected by these factors. For example, immune checkpoint inhibition (ICI) may be most effective in highly glycolytic tumors.^6^ Therefore, we set out to identify features that diminish anti-tumor immunity despite the presence of T cells and antigen hypothesizing that metabolic features diminish anti-tumor responses in an otherwise favorable TME.

To accomplish this, we began by selecting a cohort of human papillomavirus (HPV)-associated head and neck squamous cell carcinomas (HNSCs) characterized by CD8 T cell infiltration and virus-derived tumor-associated antigens.^4^ By leveraging the immune characteristics of these tumors, we hypothesized that tumor intrinsic or extrinsic features of the TME diminish the anti-tumor immune response in some, but not all tumors, focusing on metabolic features of the TME. We leveraged genomic atlases to test our hypothesis. We validated our findings in head and neck cancer across different 10 cancer types from The Cancer Genome Atlas (TCGA) and immunotherapy-treated melanomas.

## METHODS

### Clinical data collection

TCGA level 1 clinical data were abstracted from FireBrowse (http://firebrowse.org/). Data for the head and neck squamous cell carcinoma samples was derived from: gdac.broadinstitute.org_HNSCC.Merge_Clinical.Level_1.2016012800.0.0/HNSCC.clin.merged.t xt. The HPV status was identified using the variable: patient.hpv_test_results.hpv_test_result.hpv_status (levels: positive, negative, indeterminate).

### RNA sequencing and alignment

Sequencing and alignment of the TCGA data have been described previously (https://docs.gdc.cancer.gov/Data/Bioinformatics_Pipelines/Expression_mRNA_Pipeline/).

TCGA RSEM expression data were obtained through FireBrowse. File names from each cancer site are documented in **Supplemental Table S17**.

### RNA expression data preparation and analysis

RSEM expression data were extracted and pre-processed by excluding genes with zero reads across tumors. Genes with detectable reads in at least 50% of samples were included. We used the variance stabilizing transformation and normalization function in DESeq2 to normalize data for use in downstream analyses.^7^ Differential expression analysis was performed using DESeq2 on RSEM data rounded to the nearest integer. Hierarchical clustering was performed with the ComplexHeatmap R package using the “ward.D” clustering method and “pearson” distance on the rows and columns.^8^

### Single sample gene set enrichment (ssGSEA)

ssGSEA (v10.0.3) was implemented in GenePattern to estimate PA pathway and immune gene set scores (**Supplemental Table S1**).^9^ Default parameters were used with rank normalization for all ssGSEA analyses.

### Cellular abundance estimates from bulk RNAseq data using CIBERSORT

We utilized CIBERSORT in “Impute Cell Expression” mode on the TCGA transcripts per million (TPM) RNAseq data from HNSC patients to infer relative cellular proportions. For the CIBERSORT analysis, we used a head and neck reference single cell RNAseq dataset ^10^ in TPM normalization space to define proportions of tumor cells, macrophages, fibroblasts, CD8^+^ and CD4^+^ T cells in bulk RNAseq data. TCGA HNSC TPM RNAseq data were used for this analysis in order to keep the bulk RNAseq and reference matrix in the same normalization space per CIBERSORT recommendations.

### T cell status stratification

T cell infiltration scores from ssGSEA were generated using CD8^+^ T cell and cytotoxic T cell (CTL) gene sets described by Bindea et al. ^11^, **Supplemental Table S1**. We scaled ssGSEA scores from each gene set. Samples were dichotomized (T cell enriched [Thi] vs T cell depleted [Tlo]) using an upper quartile cutoff of ssGSEA scores for both CTL and CD8^+^ T cell gene sets (**Supplemental Table S2**). Tumors in the highest ssGSEA score quartile for either the CD8^+^ T cell or CTL signature were categorized as Thi and the remainder Tlo (**Supplemental Table S2**). Single sample GSEA stratification of T cell infiltration status was consistent with inferred CD8^+^ T cell abundance from computational microdissection with CIBERSORT (Wilcoxon rank-sum of CD8^+^ T cell abundance between Thi and Tlo, p < 0.001; **Supplemental Figure S1**).^12^ Single sample GSEA stratification was used for all downstream analyses for 1) comparability to other studies using the Bindea immune cell gene sets to infer immune cell responses; and 2) to use a consistent, gene set-driven approach, agnostic to the tissue of origin.

### Metabolic gene curation

We defined a set of 2,520 genes implicated in metabolism using gene sets from Broad Institute’s Molecular Signature Database (MSigDB)^13^ and Shaul *et al*.^14^

### Survival analysis

We utilized univariate Cox regression analysis to test associations between PA pathway scores and risk of mortality.

### Molecular subtype classification

To define molecular subtypes using the TCGA HNSC expression data, we utilized an R script kindly provided by the Fertig and Seiwert labs based on prior work which implements a correlation-based nearest centroid technique.^15^ Subtypes were assigned for the entire TCGA HPV+ and HPV-HNSC dataset including basal, classical, or mesenchymal subtypes.

### HPV integration

Using data from Parfenov et al., we assigned the TCGA HPV+ HNSC HPV integration status.^16^

### Tumor mutation burden

TMB data from the TCGA MC3 ^17^ were extracted using maftools^18^ We defined high TMB as ≥ 10 mutations / mega base pair (Mbp) and low TMB as < 10 mutations/Mbp.

### Single cell RNA sequencing analysis

HPV+ HNSC TIL data from Cillo *et al*^19^ were downloaded, pre-processed, normalized, and scaled using Seurat (v4.0.1).^20 21^ Data were mapped onto a single cell reference dataset to identify immune cell subsets ^20^ which were used to evaluate expression of polyamine pathway enzymes across cell types. The “AddModuleScore” function was used to determine polyamine gene set (i.e., “module”) scores using the gene sets defined below. A similar process was used for HPV-HNSC scRNAseq data from Puram *et al*^10^ which were TPM-normalized and were pre-processed, log2-transformed, and scaled.

### Statistics

R programming software (version 3.6.1) was used to perform statistical analyses.^22^ Kruskal-Wallis or Wilcoxon tests were used to compare data distributions between more than two groups or two groups, respectively, for non-normally distributed data. Chi-square tests were used to evaluate independence between groups with all expected cell counts ≥ 5. Fisher exact tests were used to test for independence with any expected cell count < 5. To account for multiple hypothesis testing, the false discovery rate was controlled using the method of Benjamini and Hochberg. Pearson correlation analyses were performed in R using the WGCNA software package.^23^ An alpha of 0.05 was used as a threshold for statistical significance, except in the case of multiple hypothesis testing where a q-value of 0.25 was used. ^*^, p ≤ 0.05; ^**^, p ≤ 0.01; ^***^, p ≤ 0.001; ^****^, p ≤ 0.0001; ns, p > 0.05.

## RESULTS

### HPV+ HNSC as a model of T cell-enriched, antigen-driven anti-tumor immunity

We stratified TCGA HPV+ HNSCs into high (Thi) and low (Tlo) T cell infiltration. Of the HPV+ HNSCs, 47% (46/97) were Thi whereas only 31% (130/420) HPV-HNSCs were Thi (χ^2^ test, p = 0.002). HPV+ HNSC patients had better survival than carcinogen-driven (HPV-) HNSCs (**Figure 1A**). Among the HPV+ HNSCs, three-year survival probability was greater for Thi (0.90, 95% CI: 0.81, 1.0) than Tlo tumors (0.55, 95% CI: 0.40, 0.75; log-rank test, p = 0.0018; **Figure 1B**). In contrast, HPV-HNSCs did not have better three-year survival when stratified by T cell status (Tlo: 0.53, 95% CI: 0.46, 0.60; Thi: 0.59, 95% CI: 0.51, 0.70; log-rank test, p = 0.42). Next, we focused our analysis on HPV+Thi HNSCs which harbor T cell infiltrates and virus-derived tumor-associated antigens.

**Figure 1.**
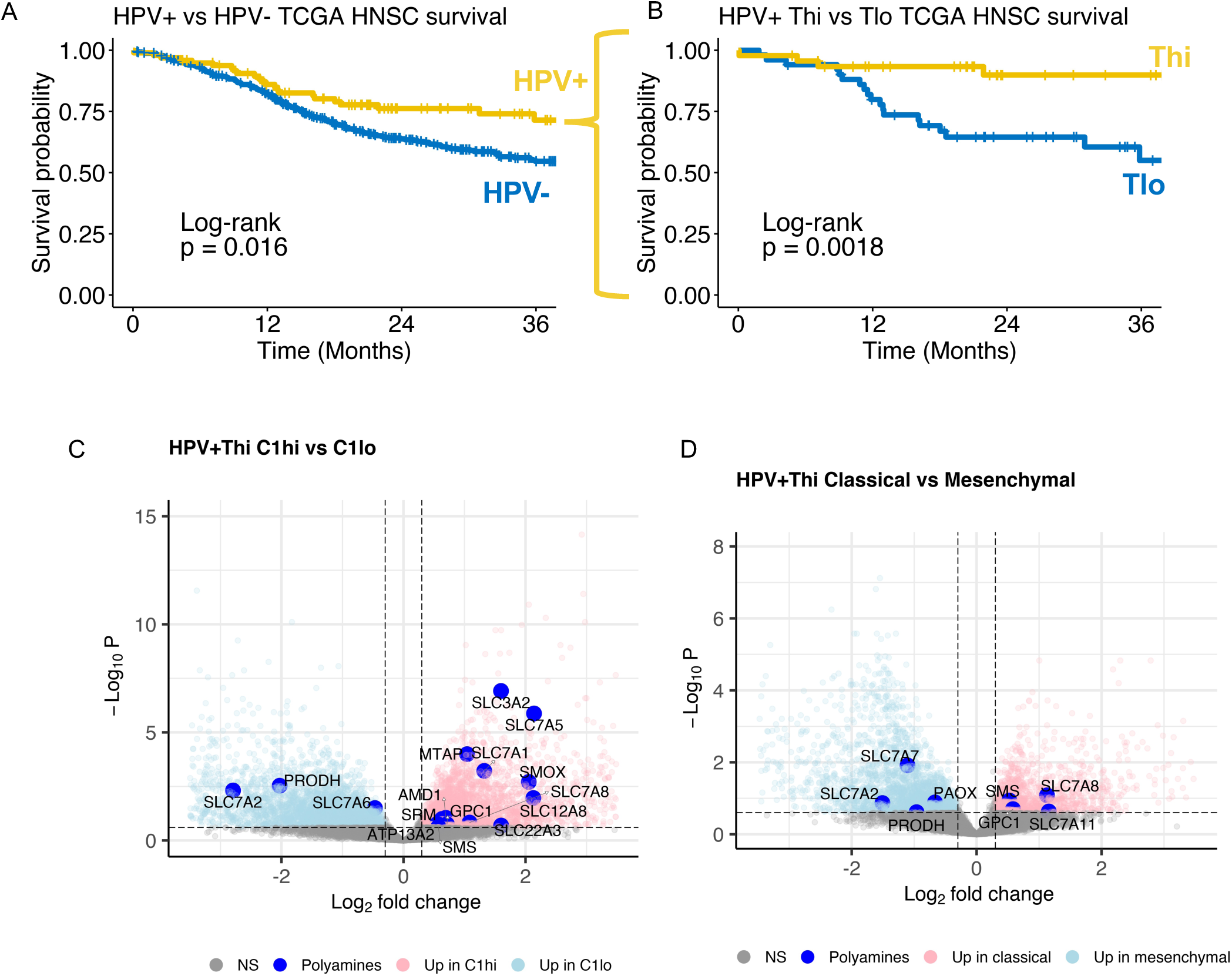
Polyamine metabolism genes are differentially expressed between T cell-infiltrated human papillomavirus (HPV)-related head and neck squamous cell carcinomas (HNSCs) stratified by prognostic molecular gene signatures. **(A)** Survival among TCGA HNSC stratified by HPV status (HPV+, n = 97; HPV-(carcinogen-driven), n = 421). **(B)** HPV+ HNSC tumors were stratified by T cell inflammation using single sample gene set enrichment (ssGSEA) Bindea *et al* CD8^+^ T cell and cytotoxic lymphocyte gene set scores. Survival based on T cell status is shown. Volcano plots demonstrate differential expression analysis results comparing (**C**) Gleber-Netto *et al* HPV+ HNSC high-risk C1 gene set high vs low among TCGA HPV+Thi HNSC and (**D**) Keck *et al* classical vs mesenchymal TCGA HPV+Thi HNSCs. *Dashed lines*, FDR q < 0.25, abs(log2 fold change) > 0.3. *Large blue points*, polyamine pathway genes.

### Differentially expressed genes (DEGs) comparing HPV+ T cell infiltrated HNSCs stratified by prognostic molecular signatures

To identify metabolic features impairing the anti-tumor immune response in an otherwise favorable TME, we performed differential expression analyses stratified by prognostic molecular phenotypes hypothesizing that metabolic features diminish anti-tumor immunity among HPV+Thi HNSCs. We focused our analysis on 2,520 metabolism-related genes derived from MSigDB (1,617 genes total) ^13 24^ and Shaul *et al* ^14^ (903 additional genes; **Supplemental Table S3**). We analyzed DE between tumors with different molecular subtypes (i.e., immune/mesenchymal vs classical) ^15^ or a high-risk HPV+ HNSC gene signature.^25^ The Keck *et al* molecular supergroups include a basal subtype characterized by a predominance of HPV-HNSCs, hypoxia, and EGFR/HER signaling.^15^ Among HPV+ HNSCs, the immune/mesenchymal subtype has a better prognosis than classical subtype tumors.^15^ The immune/mesenchymal subtype is characterized by immune markers including *CD8A, ICOS, LAG3, HLA-DRA* and mesenchymal markers such as *VIM, S100A4*, and *MMP9*. The most distinctive transcriptional feature of the classical subtype in both HPV+ and HPV-HNSCs is enrichment for polyamine catabolism gene expression.^15^ We also evaluated an HPV+ HNSC gene signature (i.e., C1) associated with high-risk HPV biology and poor prognosis.^25^

To test DE between TCGA HPV+Thi HNSCs stratified by the C1 signature, we generated a score across tumors taking the cumulative expression across C1 genes (*CDA, DFNA5, KRT14, MDFI, UPP1, DPF1, RGS20, TUBB3, PPP1R14B, RHOD, CPA4, PNLIPRP3*) for each sample and creating expression tertiles (high, intermediate, low). The distribution of C1 strata by T cell infiltration status is shown in **Table 1**.

**Table 1.** Distribution of TCGA HPV+ Thi and Tlo HNSCs by clinical and genomic features.

| Variable | T cell status |  | p-value |
| --- | --- | --- | --- |
|  | Thi, N = 46<sup>1</sup> | Tlo, N = 51<sup>1</sup> |  |
| C1 score |  |  | <0.001<sup>2</sup> |
| High | 4 / 46 (8.7%) | 28 / 51 (55%) |  |
| Intermediate | 20 / 46 (43%) | 12 / 51 (24%) |  |
| Low | 22 / 46 (48%) | 11 / 51 (22%) |  |
| Group |  |  | <0.001<sup>2</sup> |
| Basal | 0 / 46 (0%) | 15 / 51 (29%) |  |
| Classical | 8 / 46 (17%) | 30 / 51 (59%) |  |
| Mesenchymal | 38 / 46 (83%) | 6 / 51 (12%) |  |
| Site |  |  | <0.001<sup>3</sup> |
| Hypopharynx | 1 / 46 (2.2%) | 4 / 51 (7.8%) |  |
| Larynx | 2 / 46 (4.3%) | 4 / 51 (7.8%) |  |
| Oral cavity | 7 / 46 (15%) | 25 / 51 (49%) |  |
| Oropharynx | 36 / 46 (78%) | 18 / 51 (35%) |  |
| *PIK3CA* |  |  | 0.44<sup>2</sup> |
| Alteration<sup>4</sup> | 36 / 46 (78%) | 44 / 51 (86%) |  |
| No alteration | 10 / 46 (22%) | 7 / 51 (14%) |  |
| *AKT1* |  |  | 0.44<sup>2</sup> |
| Alteration<sup>4</sup> | 7 / 46 (15%) | 12 / 51 (24%) |  |
| No alteration | 39 / 46 (85%) | 39 / 51 (76%) |  |
| *MTOR* |  |  | 0.16<sup>2</sup> |
| Alteration<sup>4</sup> | 9 / 46 (20%) | 4 / 51 (7.8%) |  |
| No alteration | 37 / 46 (80%) | 47 / 51 (92%) |  |

| Variable | T cell status |  | p-value |
| --- | --- | --- | --- |
|  | Thi, N = 46<sup>1</sup> | Tlo, N = 51<sup>1</sup> |  |
| *MYC* |  |  | 0.035<sup>2</sup> |
| Alteration<sup>4</sup> | 19 / 46 (41%) | 33 / 51 (65%) |  |
| No alteration | 27 / 46 (59%) | 18 / 51 (35%) |  |

<sup>1</sup>Statistics presented: n / N (%)
<sup>2</sup>Chi-square test of independence
<sup>3</sup>Fisher's exact test
<sup>4</sup>Alteration defined a presence of mutation, copy number gain or amplification, or homozygous deletion.

Nine percent of Thi tumors and 55% of Tlo tumors were C1 high (Chi-square test, p < 0.001). DE analysis comparing HPV+Thi C1 high (C1hi; N = 4) vs C1 low (C1lo; N = 22) tumors revealed 935 metabolic genes upregulated in the HPV+Thi C1hi HNSCs (**Figure 1C, Supplemental Table S4**). Among HPV+Thi C1 high tumors, polyamine (PA) metabolism genes were upregulated (**Figure 1C**). Database for Annotation, Visualization and Integrated Discovery (DAVID) analysis demonstrated enrichment of genes involved in mitogenic signaling (*TSC2, MAPK1, INSR, PIK3CG*) plus lipid (*INPP5A, LPL*), central carbon (*IDH2, SDHB-D, HK1, HK2, LDHA, LDHC*), arginine and proline metabolism (*SMOX, ARG2, NOS2, AMD1, SRM, AGMAT, PRODH*; **Supplemental Table S5**).

Next, we compared HPV+Thi tumors based on molecular phenotype as classical versus immune/mesenchymal (**Supplemental Table S6**). Molecular subtype across TCGA HNSCs was defined using a correlation-based, nearest centroid classification approach.^15^ **Table 1** shows the distribution of molecular subtypes given T cell status across HPV+ HNSCs. Eighty-three percent of Thi tumors and 12% of Tlo tumors were classified as mesenchymal (Chi-square test, p < 0.001; **Table 1**). No Thi tumors were classified as basal. The remainder of Thi tumors were classified as classical (N = 8). DE analysis demonstrated 438 metabolic genes upregulated in the classical tumors. DAVID pathway analysis revealed upregulation of genes involved in mitogenic signaling (*GSK3B, TSC2, BRAF, PIK3CD*) in addition to lipid (*INPP4A, PLA2G2A, LPIN2*), purine (*NT5E, ENTPD1, NME3*), central carbon (*IDH1, HK2, ACAT1*), arginine and proline metabolism (*ARG2, NOS1, PRODH*; **Supplemental Table S7**). PA transport and synthesis genes were upregulated in classical tumors (**Figure 1D**). Given these findings and prior metabolomic data demonstrating enrichment of polyamines in HNSC tissues,^26 27^ we tested how polyamine expression is related to clinical (e.g., prognosis, smoking) and tumor genomic features.

### Polyamine synthesis and transport genes are associated with worse clinical and molecular features among patients with HPV+ HNSC

To characterize the extent to which PA metabolism genes are related to clinical and molecular features among HPV+ HNSCs, we performed hierarchical clustering of gene expression using a set of 35 genes involved in PA metabolism. These included polyamine metabolism genes involved in synthesis, catabolism, and transport based on literature review. Hierarchical clustering revealed three sample clusters (**Figure 2A**, *column clusters*). Oropharyngeal tumors made up most of the cohort (54/97) followed by oral cavity cancers (32/97; **Table 2**). Clusters 1 and 2 consisted of 87% and 78% oropharyngeal tumors vs Cluster 3 which was enriched in oral cavity tumors (68%; Fisher exact test, p < 0.001). Clusters 1 and 2 were predominantly Thi (69% (27/39) and 61% (11/18), respectively; **Figure 2A, Table 2**). Cluster 3 tumors were mainly Tlo (80% (32/40)). The distribution of Thi and Tlo tumors by primary site is shown in **Table 1**. Oropharyngeal tumors comprised 78% of Thi tumors and 35% of Tlo tumors. Tumor stage did not differ between clusters nor did smoking status (**Figure 2A, Table 2**).

**Figure 2.**
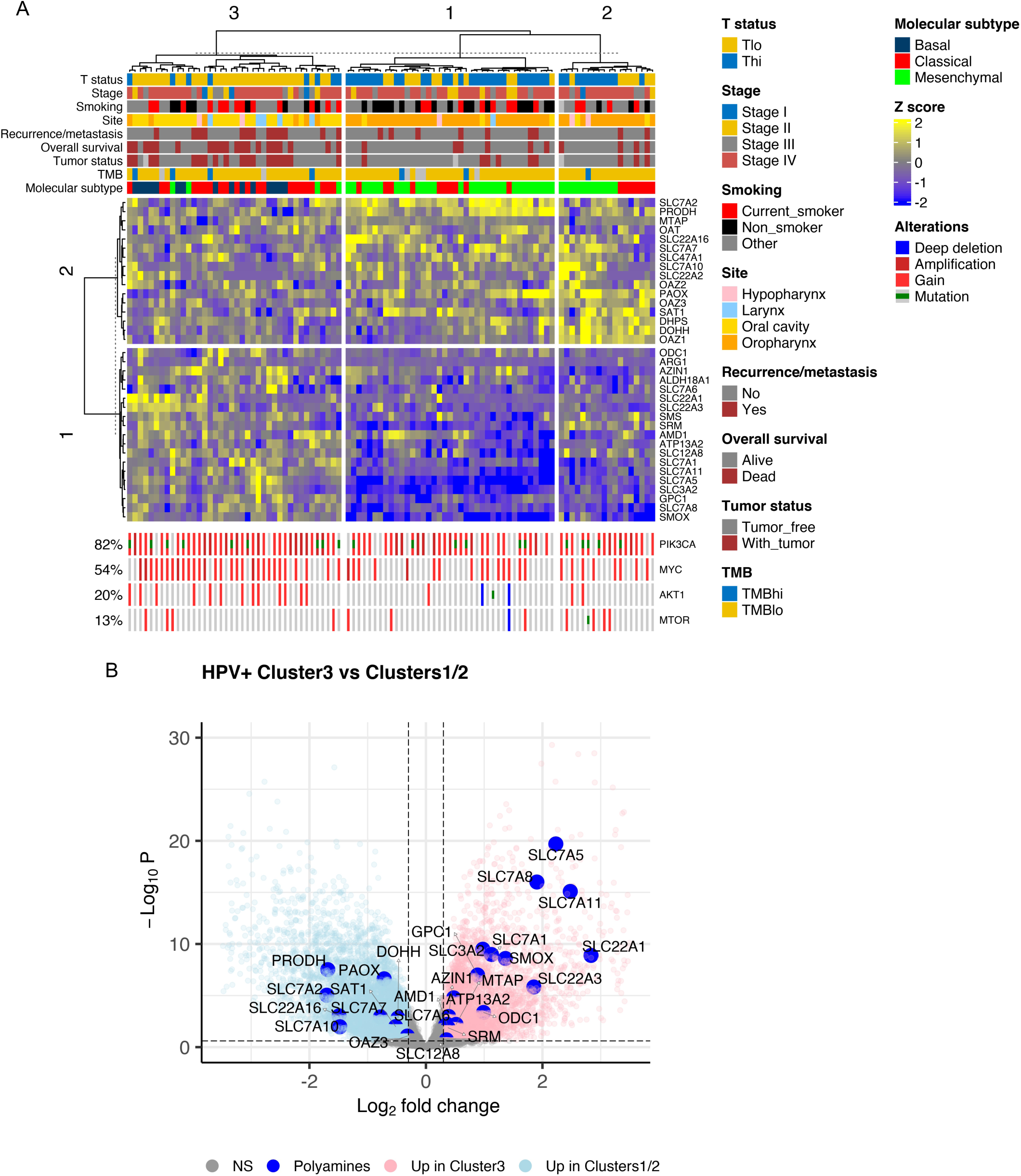
Polyamine synthesis and transport genes are associated with worse clinical and molecular features among patients with HPV+ HNSC. (**A**) Heatmap illustrates hierarchical clustering of scaled variance transformed RSEM counts for polyamine synthesis, catabolism, hypusination, and transport genes among TCGA HPV+ HNSCs (n = 97). Clinical and molecular features are annotated in the bars above the heatmap. Genomic annotations are plotted at the bottom of the heatmap for the genes shown. Rows represent unique genes. Columns represent unique patients. Hierarchical clustering was performed across columns and rows. *Stage*, AJCC 7 ^th^edition clinical stage. *TMB (tumor mutation burden)*: *TMBhi*, ≥ 10 mutations/Mbp; *TMBlo*, < 10 mutations/Mbp. (**B**) Volcano plot demonstrating differential expression analysis results comparing Cluster 3 to Clusters 1 and 2 based on the hierarchical clustering results in (**A**). *Large blue dots*, polyamine metabolism genes. *NS*, non-significant.

**Table 2.** Clinical and molecular characteristics of TCGA HPV+ HNSC clusters based on polyamine metabolism gene expression.

| Variable | Cluster |  |  | p-value<sup>2</sup> |
| --- | --- | --- | --- | --- |
|  | Cluster1, N = 39<sup>1</sup> | Cluster2, N = 18<sup>1</sup> | Cluster3, N = 40<sup>1</sup> |  |
| Tstatus |  |  |  | <0.001 |
| Thi | 27 / 39 (69%) | 11 / 18 (61%) | 8 / 40 (20%) |  |
| Tlo | 12 / 39 (31%) | 7 / 18 (39%) | 32 / 40 (80%) |  |
| Stage |  |  |  | 0.67 |
| Stage I | 0 / 39 (0%) | 1 / 18 (5.6%) | 3 / 40 (7.5%) |  |
| Stage II | 6 / 39 (15%) | 1 / 18 (5.6%) | 5 / 40 (12%) |  |
| Stage III | 6 / 39 (15%) | 3 / 18 (17%) | 7 / 40 (18%) |  |
| Stage IV | 27 / 39 (69%) | 13 / 18 (72%) | 25 / 40 (62%) |  |
| Recurrence/metastasis | 7 / 39 (18%) | 2 / 18 (11%) | 13 / 40 (32%) | 0.18 |
| Tumor status |  |  |  | 0.007 |
| Tumor free | 32 / 38 (84%) | 14 / 17 (82%) | 21 / 39 (54%) |  |
| With tumor | 6 / 38 (16%) | 3 / 17 (18%) | 18 / 39 (46%) |  |
| (Missing) | 1 | 1 | 1 |  |
| Overall survival |  |  |  | 0.003 |
| Alive | 33 / 39 (85%) | 14 / 18 (78%) | 20 / 40 (50%) |  |
| Dead | 6 / 39 (15%) | 4 / 18 (22%) | 20 / 40 (50%) |  |
| Smoking |  |  |  | 0.14 |
| Current smoker | 8 / 39 (21%) | 4 / 17 (24%) | 13 / 40 (32%) |  |
| Non-smoker | 17 / 39 (44%) | 3 / 17 (18%) | 8 / 40 (20%) |  |
| Other | 14 / 39 (36%) | 10 / 17 (59%) | 19 / 40 (48%) |  |
| (Missing) | 0 | 1 | 0 |  |
| Site |  |  |  | <0.001 |
| Hypopharynx | 1 / 39 (2.6%) | 2 / 18 (11%) | 2 / 40 (5.0%) |  |
| Larynx | 1 / 39 (2.6%) | 0 / 18 (0%) | 5 / 40 (12%) |  |
| Oral cavity | 3 / 39 (7.7%) | 2 / 18 (11%) | 27 / 40 (68%) |  |
| Oropharynx | 34 / 39 (87%) | 14 / 18 (78%) | 6 / 40 (15%) |  |
| TMB category |  |  |  | 0.88 |
| TMBhi | 2 / 35 (5.7%) | 1 / 18 (5.6%) | 4 / 38 (11%) |  |
| TMBlo | 33 / 35 (94%) | 17 / 18 (94%) | 34 / 38 (89%) |  |

| Variable | Cluster |  |  | p-value<sup>2</sup> |
| --- | --- | --- | --- | --- |
|  | Cluster1, N = 39<sup>1</sup> | Cluster2, N = 18<sup>1</sup> | Cluster3, N = 40<sup>1</sup> |  |
| (Missing) | 4 | 0 | 2 |  |
| Group |  |  |  | <0.001 |
| Basal | 0 / 39 (0%) | 0 / 18 (0%) | 15 / 40 (38%) |  |
| Classical | 10 / 39 (26%) | 7 / 18 (39%) | 21 / 40 (52%) |  |
| Mesenchymal | 29 / 39 (74%) | 11 / 18 (61%) | 4 / 40 (10%) |  |
| *PIK3CA* |  |  |  | 0.056 |
| Alteration | 28 / 39 (72%) | 15 / 18 (83%) | 37 / 40 (92%) |  |
| No alteration | 11 / 39 (28%) | 3 / 18 (17%) | 3 / 40 (7.5%) |  |
| *AKT1* |  |  |  | 0.039 |
| Alteration | 4 / 39 (10%) | 2 / 18 (11%) | 13 / 40 (32%) |  |
| No alteration | 35 / 39 (90%) | 16 / 18 (89%) | 27 / 40 (68%) |  |
| *MTOR* |  |  |  | 0.20 |
| Alteration | 4 / 39 (10%) | 5 / 18 (28%) | 4 / 40 (10%) |  |
| No alteration | 35 / 39 (90%) | 13 / 18 (72%) | 36 / 40 (90%) |  |
| *MYC* |  |  |  | 0.007 |
| Alteration | 15 / 39 (38%) | 8 / 18 (44%) | 29 / 40 (72%) |  |
| No alteration | 24 / 39 (62%) | 10 / 18 (56%) | 11 / 40 (28%) |  |

<sup>1</sup>Statistics presented: n / N (%)
<sup>2</sup>Statistical tests performed: chi-square test of independence; Fisher's exact test

R/M, tumor status, and overall survival status were worse in Cluster 3. Thirty-two percent of Cluster 3 subjects experienced R/M compared to 18% and 11% of Clusters 1 and 2, respectively (Fisher exact test, p = 0.18; **Figure 2A, Table 2**). Cluster 3 had a higher proportion of patients with recurrent or persistent tumors (“With tumor”; 46% (18/39)) than Clusters 1/2 subjects (16% (6/38) and 18% (3/17); Fisher exact test, p = 0.007; **Table 2**). Clusters 1/2 subjects experienced a lower frequency of deaths over the three-year follow-up interval (15% (6/39) and 22% (4/18), respectively) than Cluster 3 subjects (50% (20/40); Chi-square test, p = 0.003; **Table 2**). R/M was not different between clusters among Thi (Fisher exact test, p = 0.4) or Tlo subsets (Fisher exact test, p = 0.46; **Supplemental Table S8**). Thi tumors had a higher percentage of recurrent or persistent tumors in Cluster 1 and 3 (22% and 25%, respectively) compared to Cluster 2 (0%; Fisher exact test, p = 0.23; **Supplemental Table S8**). Clusters 1 and 2 exhibited upregulation of PA regulatory genes (i.e., *SAT1, OAZ1-3*; **Figure 2B**). These results suggest that PA metabolism gene expression varies by CTL infiltration and is associated with clinical features and primary site.

Next, we evaluated the relationship between tumor mutation burden (TMB) and molecular subtype with T cell infiltration among sample clusters. High TMB was present in 11% (4/38), 5.6% (1/18), and 5.7% (2/35) of tumors in Clusters 3, 2, and 1, respectively (Fisher exact test, p = 0.88; **Table 2**). Molecular subtype analysis demonstrated a high rate of mesenchymal tumors in Clusters 1 and 2 (74% (29/39) and 61% (11/18), respectively) compared to Cluster 3 which was enriched in classical subtype tumors (52% (21/40); Fisher exact test, p < 0.001; **Figure 2A, Table 2**). While the mesenchymal subtype may be associated with the degree of cancer associated fibroblasts,^10^ there was a strong association between mesenchymal subtype and Thi status (**Table 1**) suggesting that the mesenchymal signature is in part related to the extent of immune cell infiltration among these tumors.

Genomic alterations including copy number gain, amplification, homozygous deletion, and mutations of *PIK3CA, AKT1, MTOR*, and *MYC* were analyzed given their role in regulating PA metabolism. *AKT1* and *MYC* were altered more frequently in Cluster 3 (32% and 72%, respectively) compared to Clusters 1 and 2 (10% and 38% vs 11% and 44%, respectively; *AKT1*: Fisher exact test, p = 0.039; *MYC*: Chi square test, p = 0.007; **Table 2**). *PIK3CA* was altered in 72%, 83%, and 92% of cases in Clusters 1, 2, and 3, respectively (Fisher exact test, p = 0.056; **Table 2**). *MYC* alterations were more frequent among the Tlo Cluster 3 samples (78%) compared to Clusters 1 and 2 (33% and 57% respectively; Fisher exact test, p = 0.019), but there was no difference in *MYC* alterations between clusters in the Thi tumors (Fisher exact test, p = 0.84; **Supplemental Table S8**). *MTOR* alterations were more frequent among Thi tumors in Clusters 2 (45%) and 3 (25%) compared to Cluster 1 (7%; Fisher exact test, p = 0.016), but there was no difference in *MTOR* alterations in the Tlo tumors (Fisher exact test, p = 0.31; **Supplemental Table S8**). *PIK3CA* and *AKT1* did not vary between clusters. Genomic alterations did not vary by T cell status, except for *MYC* which was enriched in Tlo tumors (65% vs 41% of Thi tumors; Chi-square test, p = 0.035; **Table 1**). These data provide evidence that specific genomic and molecular features of HPV+ HNSCs are associated with PA metabolism gene expression in a CTL infiltration-dependent manner.

Genewise clustering analysis was also performed. PA genes clustered into two groups. Group 1 included genes involved in synthesis (*ODC1, ARG1, AZIN1, SMS, SRM, AMD1, GPC1*) and transport, and Group 2 included genes involved in catabolism (*OAZ1, OAZ2, OAZ3, SAT1*), hypusination (*DOHH, DHPS*), and transport (**Figure 2A**). Group 1 PA genes were differentially enriched in Cluster 3 tumors (except *ARG1, ALDH18A1*, and *SMS*) and downregulated in Clusters 1 and 2 (**Figure 2B, Supplemental Table S9**). In comparison, PA synthesis genes (*SRM, AMD1, ARG1, ALDH18A1*), transporters (e.g., *SLC3A2* and *GPC1*), and *SMOX* were enriched in HPV+Tlo relative to HPV+Thi HNSCs (**Supplemental Figure S2A, Supplemental Table S10**). HPV-HNSCs were also differentially enriched in PA synthesis genes (*SRM, ODC1*), transporters (*SLC3A2, GPC1, SLC7A1*), and *SMOX* expression compared to HPV+ HNSC (**Supplemental Figure S2B, Supplemental Table S10**). Thus, our next objective was to evaluate tumor-intrinsic and -extrinsic sources of PA pathway gene expression.

### PA gene expression and tumor intrinsic features

To evaluate the relationship between pathway-level gene expression and tumor-intrinsic features, we defined four PA gene sets (synthesis, catabolism, transport, and combined; **Figure 3**). These gene sets were generated in part based on prior work.^28^ The combined gene set was generated using PA genes that were negatively correlated with both Bindea cytotoxic lymphocyte gene set (CTL) and REACTOME IFN_Y_ gene set ssGSEA scores across multiple TCGA T cell-infiltrated tumor types (**Supplemental Figures S3A and B**).^29^ *SMOX* and *PAOX* were included with the synthetic genes as they regenerate spermidine (*SMOX, PAOX*) and putrescine (*PAOX*). *ALDH18A1* and *PRODH* were included as they catalyze synthesis of pyrroline-5-carboxylate from glutamate and proline, respectively, which are converted to ornithine via *OAT* (ornithine aminotransferase). The transporters included have been previously reported as putative polyamine transporters.^28 30-41^

**Figure 3.**
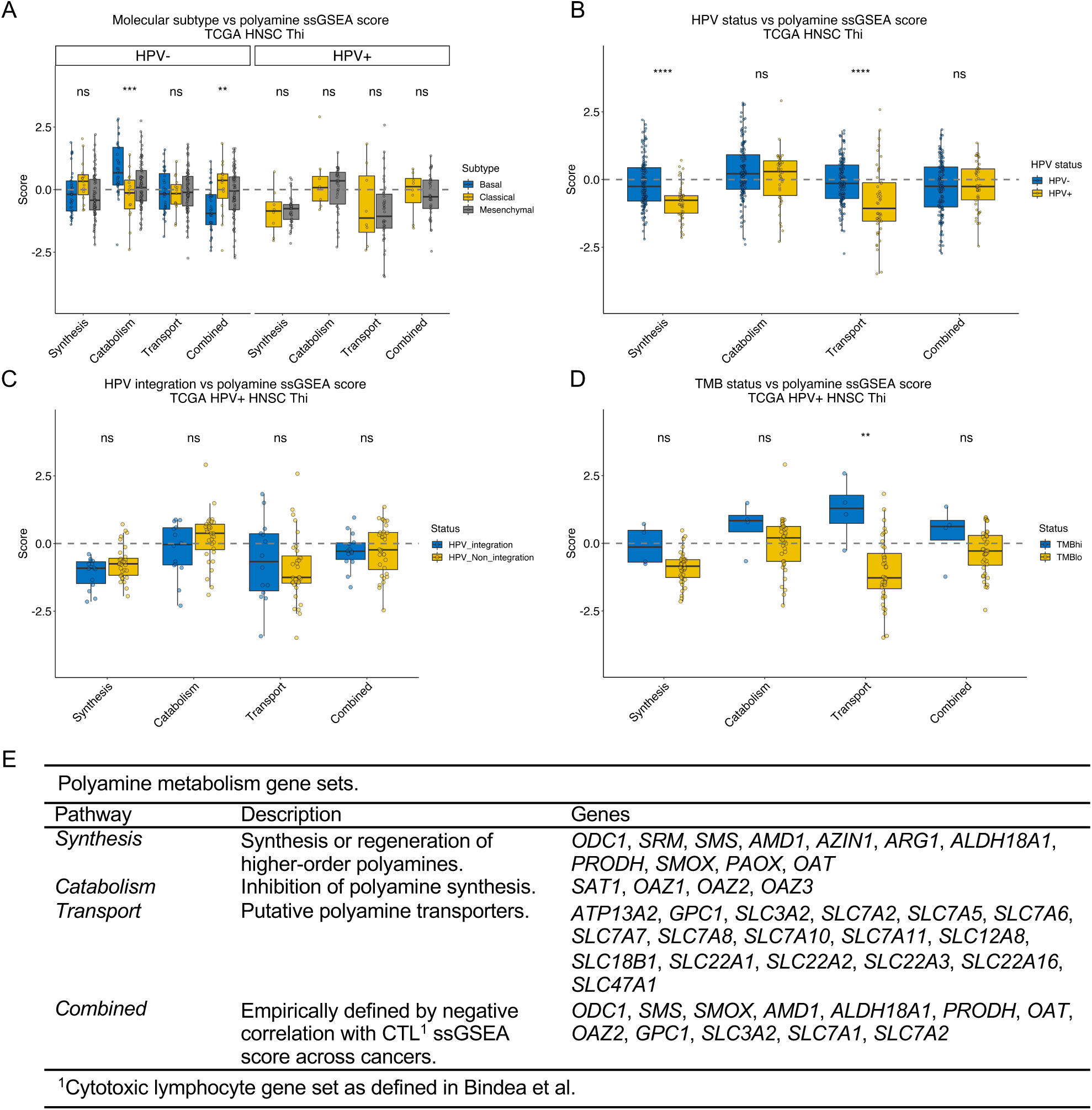
Polyamine metabolism gene expression and tumor-intrinsic features. Polyamine gene expression was grouped by functional category into synthesis, catabolism, transport, and combined gene sets (as in **Table 6**) and single sample gene set enrichment analysis (ssGSEA) was performed to generate pathway scores. Polyamine gene set scores across TCGA Thi HNSCs were stratified by **(A)** molecular subtype, **(B)** HPV status, **(C)** HPV integration status as defined in Parfenov *et al*, and **(D)** tumor mutation burden (*TMBhi*, ≥ 10 mutations/Mbp; *TMBlo*, < 10 mutations/Mbp). **(E)** Polyamine metabolism gene sets. ^**^, p ≤ 0.01; ^***^, p ≤ 0.001; ^****^, p ≤ 0.0001; ns, p > 0.05.

We hypothesized that PA metabolism genes vary by molecular subtype. There were no differences in PA ssGSEA scores based on molecular subtype among HPV+ HNSCs (**Figure 3A**). HPV-HNSCs exhibited higher catabolism and lower combined PA ssGSEA scores among the basal phenotype without differences in synthesis or transport. HPV+Tlo HNSCs also lacked differences in PA ssGSEA scores except for transport which was significantly higher among basal tumors (**Supplemental Figure S4**).

We also characterized PA pathway expression based on HPV status. Prior studies identified that viral replication depends on PAs ^42^ and that some viruses may stimulate PA synthesis.^43 44^ There are also data demonstrating nicotine-driven *ODC1* expression.^45^ PA synthesis and transport ssGSEA scores were higher among HPV-Thi versus HPV+Thi HNSCs (**Figure 3B**). PA synthesis was higher, and catabolism was lower among HPV-smokers compared to non-smokers (**Supplemental Figure S5**). HPV+ smokers demonstrated a non-significant trend towards higher PA synthesis than non-smokers (**Supplemental Figure S5**).

Next, we examined whether HPV integration impacts PA pathway expression. We evaluated the association between PA pathway ssGSEA scores and HPV integration.^16^ Defining tumors as HPV-integrated or HPV-non-integrated, we observed no statistically significant differences in PA pathway expression based on integration status in Thi or Tlo HPV+ HNSCs (**Figure 3C, Supplemental Figure S4C**).

Lastly, we tested the relationship between TMB and PA gene set expression given that PAs are involved in epigenetic regulation and DNA stabilization. Among HPV+Thi HNSCs, there was no difference in PA synthesis, catabolism, or combined ssGSEA scores based on TMB (**Figure 3D**). However, PA transport scores were higher in the TMBhi tumors. Collectively, these data demonstrate that integration status, TMB, or molecular phenotype do not fully account for the variance in PA expression among HPV+Thi HNSCs; although, HPV status and smoking may influence PA metabolism.

### PA pathway expression and TME cellular composition

Given the importance of PA metabolism in immune cell function, we assessed PA expression in the TME. With bulk RNAseq data from TCGA HPV+Thi HNSCs, CIBERSORT was used to infer the cellular composition of the TME including tumor cells, macrophages, CD8^+^ and CD4^+^ T cells, and fibroblasts. These data were used to test the correlation between cell type abundance and ssGSEA scores for PA pathway expression (**Figure 4**). We observed non-significant negative correlations between macrophage abundance and PA synthesis (R = -0.28, p = 0.061; **Figure 4A**) and significant positive correlations with catabolism and transport (R = 0.35, p = 0.016; R = 0.32, p = 0.032; **Figures 4B**,**C**). Tumor cell abundance was positively correlated with synthesis though failed to reject the null-hypothesis (R = 0.29, p = 0.054; **Figure 4A**) and negative correlated with transport (R = -0.36, p = 0.015; **Figure 4C**). Fibroblast abundance was negatively correlated with catabolism (R = -0.41, p = 0.0045; **Figure 4B**). CD4^+^ and CD8^+^ T cell abundance was not correlated with PA pathway ssGSEA scores except for CD4^+^ T cells which were positively correlated with the combined PA pathway (R = 0.39, p = 0.0075; **Figure 4D**). We also analyzed the synthesis:catabolism, synthesis:transport, and catabolism:transport ratios to assess the magnitude of the relationship between PA gene set scores and cellular abundance (**Supplemental Figure S6**). The strongest relationship was observed between macrophage abundance and the synthesis:catabolism ratio strongly favoring catabolism (R = -0.45, p = 0.0017; **Supplemental Figure S6**).

**Figure 4.**
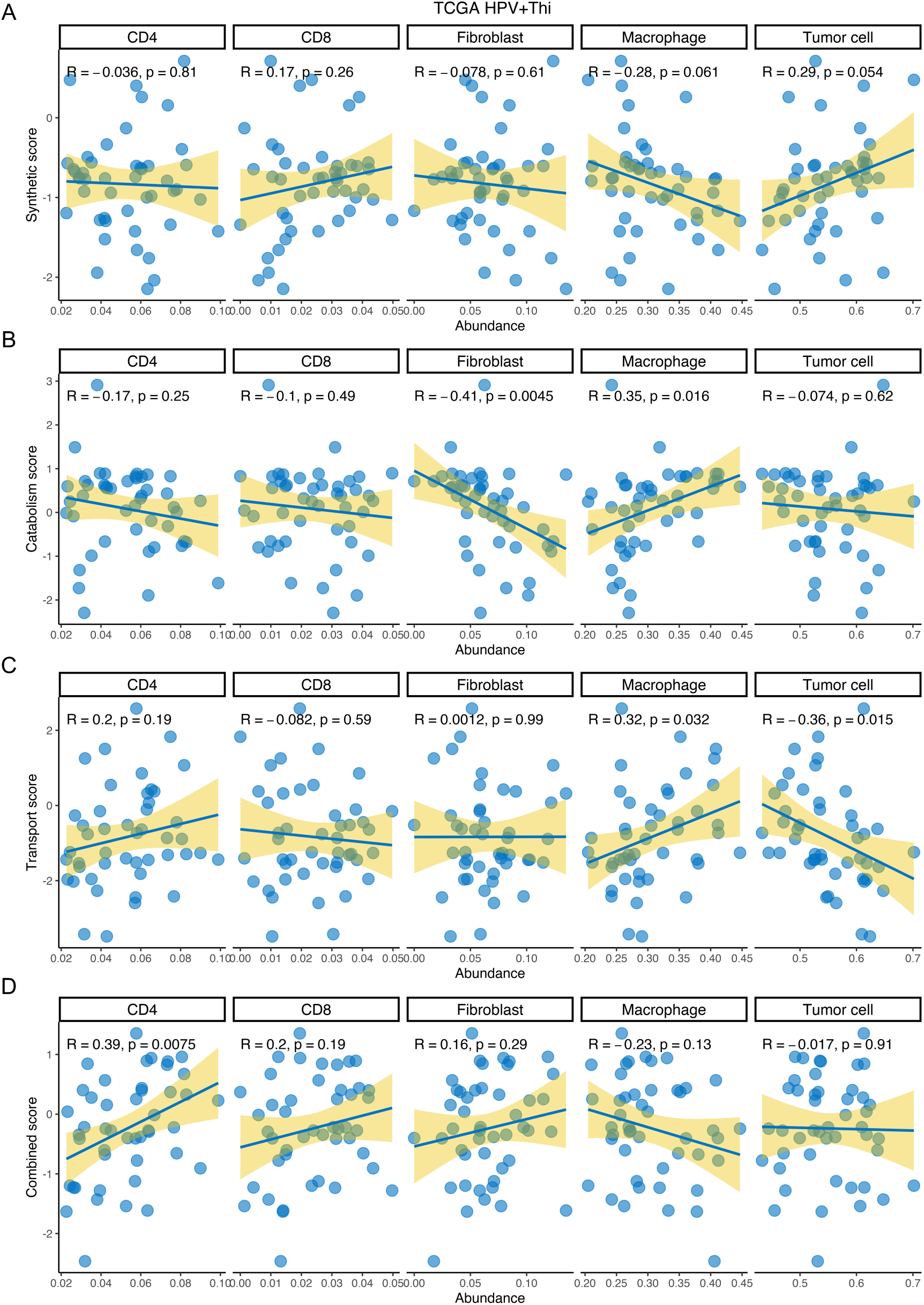
Polyamine gene expression and cellular abundance in HPV+Thi HNSC. CIBERSORT was used to define cellular abundance as a proportion of cell types in the tumor microenvironment using TCGA HPV+Thi HNSCs. Puram *et al* single cell signature matrix was used to define cell lineages. Cellular fractions were stratified by polyamine **(A)** synthesis, **(B)** catabolism, **(C)** transport, and **(D)** combined pathway ssGSEA scores. Relative cellular abundance is plotted on the x-axis and polyamine ssGSEA scores are plotted on the y-axis. Pearson correlation coefficients and p-values are displayed in the plot windows.

### PA gene expression among HNSC TILs

Given the importance of PA expression to T cell and macrophage function,^46 47^ we utilized single cell RNAseq (scRNAseq) data from Cillo *et al* to examine PA expression across HPV+ HNSC TILs (**Figure 5**).^19^ We generated single cell pathway scores using the PA gene sets defined above which were projected onto single cells (**Figure 5A-C**). In agreement with the bulk RNAseq data, PA catabolism was significantly enriched in CD16^+^ and CD14^+^ monocytes relative to other lineages (**Supplemental Tables S12**,**13**). PA catabolism pathway expression was differentially enriched among CD16^+^ and CD14^+^ monocytes and dendritic cells (**Supplemental Figure S7A**; **Supplemental Table S14**). In contrast, the PA synthesis pathway gene set was highly expressed in Tregs, and the PA transport pathway gene set was most highly expressed among CD4^+^ CTLs (**Figure 5**; **Supplemental Table S12-13**). Moreover, analysis of scRNAseq data from 21 HPV-HNSCs^10^ demonstrated greater PA catabolism module expression in macrophages compared to T cells or tumors cells (**Supplemental Figure S8; Supplemental Tables S15 and S16**). Moreover, HPV-tumor cells had greater transport and combined pathway gene set expression than T cells, macrophages, or fibroblasts and higher synthesis pathway gene set expression than T cells or fibroblasts (**Supplemental Figure S8; Supplemental Tables S15-16**). The scRNAseq data suggest that myeloid cells upregulate PA catabolism genes to a greater extent than other cells of the TME, consistent with the bulk RNAseq data.

**Figure 5.**
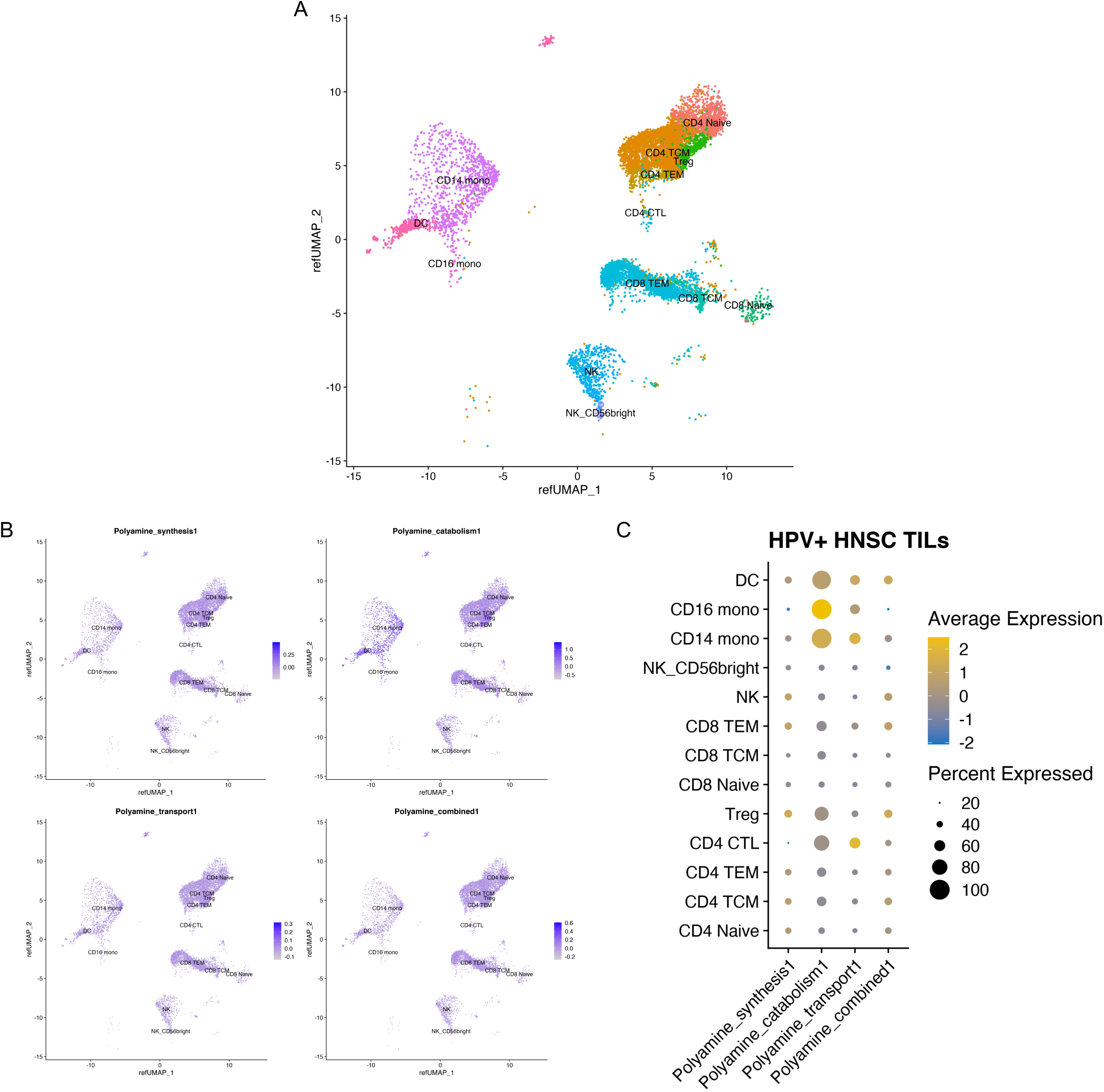
Polyamine pathway expression among tumor infiltrating lymphocytes (TIL) in HPV+ HNSC. TIL single cell RNA sequencing data (Cillo *et al*) from eight HPV+ HNSCs (14,859 cells) were mapped onto a reference single cell data set to infer cell lineage. (**A**) UMAP demonstrating lineage assignments across HPV+ HNSC TILs. (**B**) Polyamine metabolism pathway expression overlaid onto UMAP demonstrating pathway expression among clusters and quantified in (**C**).

### Pan-cancer polyamine gene set expression and immune response

We tested the relationship between PA pathway gene set expression and TCR clonality.^48^ We generated PA synthesis, catabolism, transport, and combined pathway gene set tertiles and compared scaled Richness and Shannon scores between gene set strata among TCGA HPV+ HNSCs. TCR richness was negatively associated with PA synthesis (Kruskal-Wallis test, p = 0.008; **Supplemental Figure S9**). Shannon entropy demonstrated a similar negative relationship with PA synthesis (Kruskal-Wallis test, p = 0.007; **Supplemental Figure S9**). Other PA pathway gene set scores were not associated with TCR clonality among these tumors (**Supplemental Figure S9**).

Next, we assessed the relationship between PA pathway gene set expression with chemokine and immune checkpoint gene expression across cancer types in TCGA tumors. We defined tumors with high T cell infiltration across 10 cancer types (BLCA, LUSC, LUAD, HNSC, CESC, BRCA, KIRC, THCA, SKCM, PRAD) as well as three subtypes (HPV+ CESC, HPV+ HNSC, and HPV-HNSC; too few HPV-CESC for analysis) using the ssGSEA approach as described above. Using these Thi data sets, we performed correlation analyses comparing PA gene set ssGSEA scores and expression of genes involved in the recruitment of effector CD8^+^ T cells (*CXCL9, CXCL10, CXCL11, IRF1*) or Tregs (*CCL17, CCL22;* **Supplemental Figure S10**). *CXCL9, CXCL10, CXCL11, IRF1* were negatively correlated with the PA combined ssGSEA scores in four (R range: -0.31, -0.15), four (R range: -0.22, -0.15), three (R range: -0.23, -0.15), and three (R range: -0.32, -0.21) cancer types, respectively (FDR q < 0.25; **Supplemental Figure S10**). *CXCL9, CXCL10, CXCL11, IRF1* were negatively correlated with the PA transport ssGSEA scores in two (R range: -0.28, -0.22), four (R range: -0.33, -0.26), five (R range: -0.32, - 0.13), and six (R range: -0.25, -0.16) cancer types, respectively (FDR q < 0.25; **Supplemental Figure S10**). *CXCL9, CXCL10, CXCL11, IRF1* were positively correlated with the PA catabolism ssGSEA scores in one (R: 0.15), six (R range: 0.14, 0.35), six (R range: 0.15, 0.36), and six (R range: 0.15, 0.32) cancer types, respectively (FDR q < 0.25; **Supplemental Figure S10**). *CCL17* was negatively correlated with the PA combined ssGSEA scores in seven cancer types (R range: -0.33, -0.15; FDR q < 0.25) and had few positive correlations with any of the PA gene sets. *CCL22* was positively correlated with the HPV+ and HPV-HNSC and SKCM PA transport ssGSEA scores but had few correlations with other PA pathway scores (R range: 0.25, 0.39; FDR q < 0.25).

PA catabolism and transport pathway scores were positively correlated with markers of T cell activation or exhaustion. Catabolism gene set scores were positively associated with *PDCD1* expression in five (R range: 0.20, 0.28), *HAVCR2* in seven (R range: 0.19, 0.39), *LAG3* in seven (R range: 0.15, 0.44), and *PDCD1LG2* in six cancer types, respectively (R range: 0.18, 0.35; FDR q < 0.25; **Supplemental Figure S11**). Transport scores were positively associated with *HAVCR2* in eight *(R range: 0*.*12, 0*.*44)* and *PDCD1LG2* in seven cancer types, respectively (R range: 0.11, 0.52; FDR q < 0.25; **Supplemental Figure S11**). The PA combined scores were negatively correlated with *PDCD1* in 10 cancer types (R range: -0.32, -0.13), *HAVCR2* in nine (R range: -0.32, -0.11), *TIGIT* in six (R range: -0.40, -0.09), *LAG3* in eight (R range: -0.38, - 0.25), *CD274* in five (R range: -0.29, -0.11), and *PDCD1LG2* in eight cancer types (R range: - 0.37, -0.12; FDR q < 0.25; **Supplemental Figure S11**).

Correlation analyses were performed between the REACTOME IFN*γ* signaling gene set or the Bindea CTL ssGSEA scores and PA pathway ssGSEA scores (**Figure 6A**). The IFN*γ* ssGSEA scores were negatively correlated with PA combined and synthesis ssGSEA scores in seven (R range: -0.42, -0.17) and seven (R range: -0.33, -0.19) cancer types, respectively (FDR q < 0.25). IFN*γ* scores were positively correlated with catabolism scores in four cancer types.

**Figure 6.**
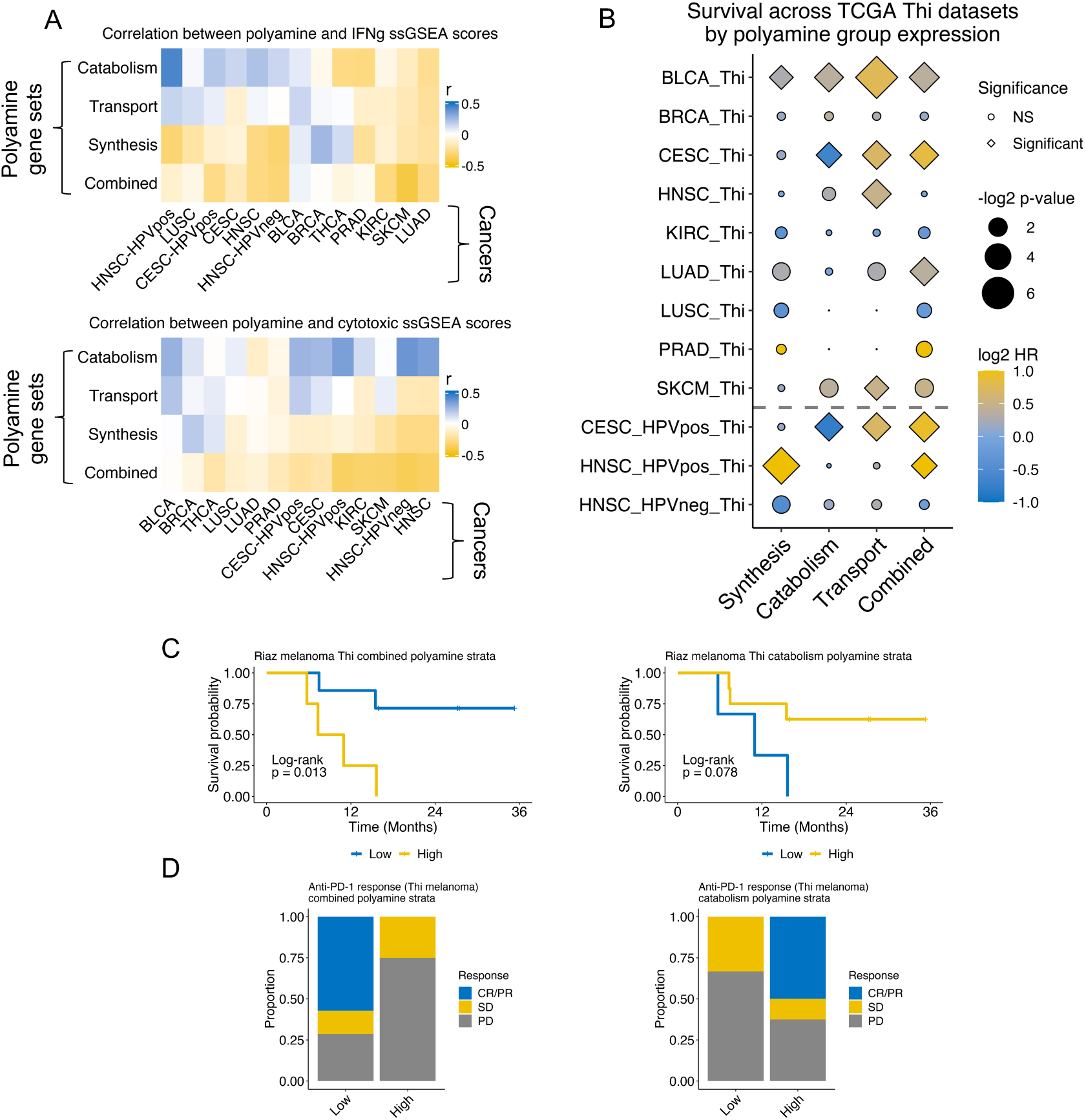
Immune functional markers and survival vary with polyamine pathway ssGSEA scores across T cell-inflamed cancers. Ten of the mostly highly inflamed cancer types were stratified by T cell infiltration status (high vs low) using TCGA bulk RNAseq data as described before. **(A)** Correlation between polyamine ssGSEA scores and REACTOME IFN*γ* pathway (*upper*) and Bindea *et al* cytotoxic lymphocyte pathway (*lower*) ssGSEA scores are shown in the heatmaps. Each rectangle represents a correlation coefficient for the correlation between polyamine ssGSEA score and immune pathway ssGSEA score for each cancer type. **(B)** Pan-cancer Cox regression analysis among the Thi tumors for each cancer type was performed using continuous polyamine ssGSEA scores as a covariate. Log2 hazard ratios (HR) are plotted. Diamonds represent statistically significant associations (*Log test -log2 p-value*, FDR q < 0.25) and circles represent non-significant associations; size of shape represents magnitude of the q-value. **(C)** Survival among Nivolumab-treated Thi melanomas (n = 11; Riaz *et al*) by combined (*left panel*; high: n = 4, low: n = 7) or catabolic (*right panel*; high: n = 8, low: n = 3) polyamine ssGSEA score strata. (**D**) Barplots showing proportion of complete or partial responders (CR/PR), stable disease (SD), or progressive disease (PD) given combined (*left panel*) or catabolic (*right panel*) polyamine ssGSEA score strata.

Similarly, CTL gene set scores were negatively correlated with PA combined pathway scores in 10 cancer types (R range: -0.39, -0.12) and positively correlated with PA catabolism scores in 10 cancer types (R range: 0.09, 0.36; FDR q < 0.25; **Figure 6A**). We next sought to characterize the association between PA pathway ssGSEA scores and survival across cancers.

### Pan-cancer polyamine gene set expression and outcomes in T cell-enriched tumors

We performed Cox regression testing the association between PA pathway ssGSEA scores and risk of mortality. High transport scores were associated with a greater risk of mortality among BLCA, CESC, HNSC, LUAD, SKCM, and HPV+ CESC cohorts. Combined PA pathway gene set scores were associated with higher mortality among the BLCA, CESC, LUAD, HPV+ CESC, and HPV+ HNSC cohorts, and a trend in worse survival among the PRAD and SKCM data sets (**Figure 6B**). In contrast, PA combined pathway gene set scores were not associated with risk of mortality across Tlo tumors (**Supplemental Figure S12**). However, higher PA transport scores were associated with worse survival among Tlo CESC, SKCM, and BLCAs.

Given the negative association between the PA combined pathway ssGSEA scores with effector lymphocyte gene set scores and prognosis, we hypothesized that PA expression affects response to ICI. We evaluated the relationship between PA combined and catabolism pathway scores with survival among a subset of T cell-enriched, Nivolumab-treated melanomas. ^49^ High combined PA pathway ssGSEA scores were associated with worse survival while the opposite was true based on catabolism pathway ssGSEA scores (**Figure 6C**). Response to ICI tracked with the survival data for the combined and catabolism pathway scores (**Figure 6D**).

## DISCUSSION

In this study, we leveraged T cell-infiltrated, immunogenic, HPV-associated head and neck cancer as a model for identifying metabolic features associated with decreased anti-tumor immunity. Our approach offered the opportunity to evaluate tumor and stromal programs involved in diminishing the effects of CTLs. Using this strategy, polyamine synthesis and transport gene expression were linked with aggressive molecular phenotypes, lower anti-tumor immunity, poor prognosis across cancer types, and a poor response to immunotherapy among melanomas.

Polyamines are a family of low molecular weight polycations that regulate multiple cell processes from proliferation and adaptive immunity^50 51^ to epigenetic modifications,^52 53^ metabolite availability,^54 55^ transcriptional regulation,^56^ and chromatin stabilization.^57^ Cells primarily synthesize PAs but can also acquire them from the TME. Given the essential role of PAs in tumor and T cell function,^46^ this pathway may be rationally targeted to diminish tumor growth and/or enhance anti-tumor immunity.

Dysregulation of the TME metabolic milieu tamps anti-tumor immune control, in part, by producing inhibitory metabolites, depleting essential nutrients, and creating a hostile hypoxic and acidic ecosystem. In our analysis, polyamine gene expression was upregulated in aggressive molecular phenotypes among HPV-related HNSC tumors with abundant T cells and antigen. Notably, some T cell infiltrated HPV+ HNSCs clustered with T cell-depleted HPV+ HNSCs sharing upregulation of genes involved in polyamine synthesis and transport. Clusters predominantly containing T cell-infiltrated tumors had upregulation of genes involved in polyamine catabolism and regulation (i.e., *SAT1, OAZ1*-3). However, using bulk RNAseq, it was unclear whether these expression patterns were related to tumor-intrinsic or -extrinsic features.

Analyzing polyamine gene expression and tumor-intrinsic molecular features, polyamine expression varied by HPV status, but not integration. HPV+Thi HNSCs with high TMB were associated with high polyamine transport gene set scores. At the single cell level, PA catabolism and transport pathway gene set scores were enriched in the myeloid compartment of HPV+ HNSCs. Whether this is directly or indirectly associated with T cell dysfunction is unclear. In the pan-cancer analysis, we observed negative correlations between polyamine synthesis or combined pathway scores with effector lymphocyte scores. Moreover, combined pathway scores were associated with poor survival across several cancers and poor response to anti-PD-1 therapy. Taken together, these results suggest prognostic and immunotherapeutic implications for PA metabolism.

In this study, we identified worse survival across cancer types in tumors with high combined PA pathway scores suggesting that polyamine synthesis and transport blockade may benefit cancer patients in a T cell-dependent manner. Polyamine blockade therapy (AMXT1501/DFMO) using difluoromethylornithine (DFMO), an ornithine decarboxylase inhibitor, and AMXT 1501,^28 58^ a polyamine transport inhibitor, demonstrates excellent responses in preclinical models of diffuse intrinsic pontine glioma ^59^ or *MYCN* transgenic mice.^28^ AMXT1501/DFMO shows activity in immunocompetent, but not immunodeficient mouse tumor models.^60^ In contrast, either agent alone does not provide equivalent responses to the combined therapy, likely secondary to compensatory mechanisms. This may account for the limited responses noted to date with polyamine monotherapy.^61 62^ In this regard, these data identify potential biomarkers for patient selection in employing agents of the PA pathway such as in the case of HPV+ HNSC patients with increased T cell infiltrates and high polyamine levels.

PA metabolism is essential for immune cell function. Several metabolites contribute to PA synthesis including methionine, glutamine, arginine, and proline. About 30% of polyamines are derived from glutamine in activated T cells, though polyamine synthesis blockade prevents T cell expansion.^46^ In macrophages, inhibition of either PA synthesis or hypusination diminishes oxidative phosphorylation and prevents alternative macrophage differentiation. ^47^ Disruption of PA homeostasis in the TME may affect the function of infiltrating immune cells. One possibility is that tumor cell death exposes cells in the TME to millimolar intracellular concentrations of PAs which could promote an immunosuppressive immune infiltrate. Another possibility is that tumors leverage PAs for intrinsically-mediated immune evasion by inducing autophagy, ^63^ favorable epigenetic alterations, or stabilizing their DNA from the effects of cytotoxic lymphocytes. ^57^ These questions warrant further research.

Future studies will need to address some of the limitations of this study including the small sample size of HPV+ HNSCs, limiting our ability to draw conclusions especially in terms of survival and patient outcomes. Larger cohorts powered to detect differences in survival and/or treatment outcomes as a function of metabolic perturbations is the next step in defining mechanisms that affect anti-tumor immunity. Studies defining the mechanism and extent to which PA metabolism diminishes the anti-tumor immune response are also needed. Whether this occurs because of exposing the TME to millimolar intracellular PAs from tumor cell death or through cell-intrinsic mechanisms that promote tumor cell survival, or some combination of these possibilities remains unknown. Lastly, the role of PA transport in tumor cell proliferation and T cell function is unknown. The degree to which putative PA transporters regulate nutrient transport and which transporters play a dominant role in PA metabolism requires further investigation. This could yield fundamental insight into differential regulation of metabolites in the TME and ensuing anti-tumor immune responses.

## Data Availability

TCGA expression data are available from FireBrowse (http://firebrowse.org/). Accession numbers for data accessed through the NCBI Gene Expression Omnibus include GSE91061, GSE10322, and GSE139324.

## DECLARATIONS

### Ethics Approval and Consent to Participate

Data were obtained from publicly available databases.

### Consent for Publication

Not required

### Availability of Data and Material

TCGA expression data are available from FireBrowse (http://firebrowse.org/) with the filenames defined in **Supplemental Table S17**. Accession numbers for data accessed through the NCBI Gene Expression Omnibus include GSE91061,^49^ GSE10322,^10^ and GSE139324.^19^ Source code is available online through the GitHub repository: https://github.com/alexharbison/polyamines_immunometabolism_cancer.git (DOI: 10.5281/zenodo.4959622).

### Authors Contributions

R.A.H., T.S., C.F., E.F., and J.P. conceived the study. R.A.H. and R.E. collected the data. R.A.H. analyzed the data. R.A.H. and M.C. generated and revised source code. R.A.H., R.P., M.C., R.L., M.B., T.M-S., R.C., T.S., C.F., E.F., and J.P. interpreted the data. R.A.H., E.F., and J.P. drafted the article. All authors participated in critical revision of the article. R.A.H., E.F., and J.P. were involved in final approval of the article.

## Acknowledgements

The authors would like to thank the members of the Powell lab for their thoughtful feedback and advice. We would like to acknowledge Ludmila Danilova for critical feedback regarding the manuscript.

## LIST OF ABREVIATIONS

CTL: cytotoxic lymphocyte
GSEA: gene set enrichment analysis
HNSC: head and neck squamous cell carcinoma
HPV: human papillomavirus
ICI: Immune checkpoint inhibition
PA: polyamine
R/M: recurrent/metastatic
ssGSEA: single sample gene set enrichment analysis
TCGA: The Cancer Genome Atlas
Thi: T cell enriched
TIL: tumor infiltrating lymphocytes
Tlo: T cell depleted
TMB: tumor mutation burden

