## Supplemental Figures for "Interrogation of T Cell-Enriched Tumors Reveals Prognostic and Immunotherapeutic Implications of Polyamine Metabolism"

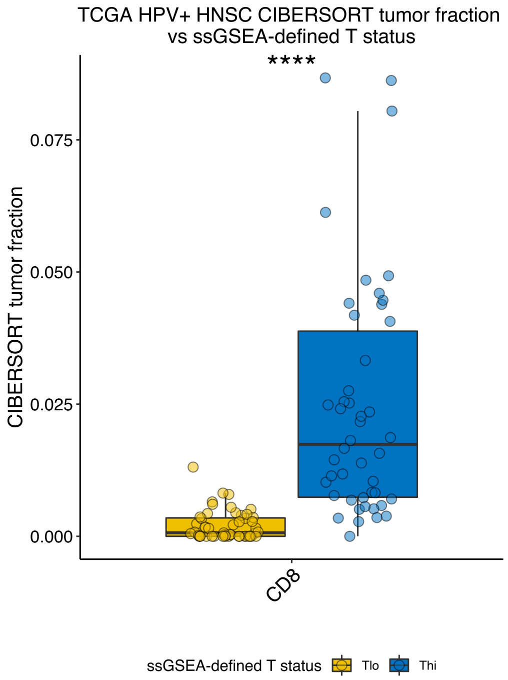


**Figure S1. CIBERSORT relative CD8+ T cell tumor fraction versus ssGSEA-defined T cell infiltration status.** CD8+ T cell infiltration as a proportion of cells in the tumor microenvironment inferred from bulk RNAseq data using CIBERSORT is stratified by ssGSEA-defined T cell infiltration status among HPV+ HNSCs. ****, $p \leq 0.0001$.

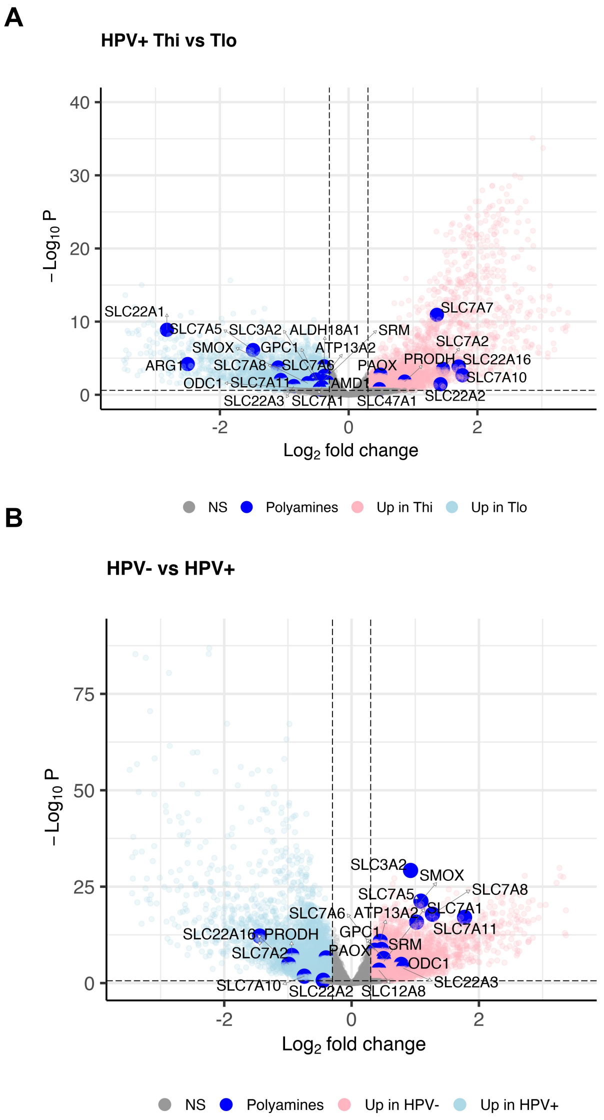


**Figure S2. Differential expression analyses of TCGA HNSC RNAseq data.** (**A**) HPV+ Thi vs Tlo HNSC. *Pink circles*, gene upregulated in Thi tumors. *Light blue circles*, genes upregulated in Tlo tumors. (**B**) HPV+ vs HPV- HNSC. *Pink circles*, gene upregulated in HPV- tumors. *Light blue circles*, genes upregulated in HPV+ tumors. *Dark blue circles*, polyamine pathway genes labeled.

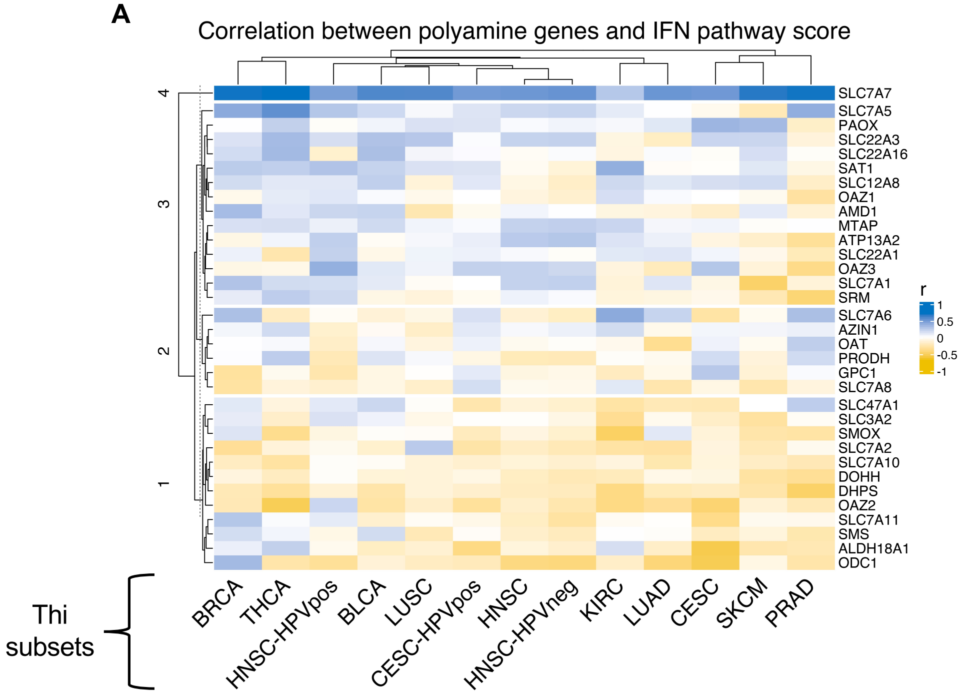


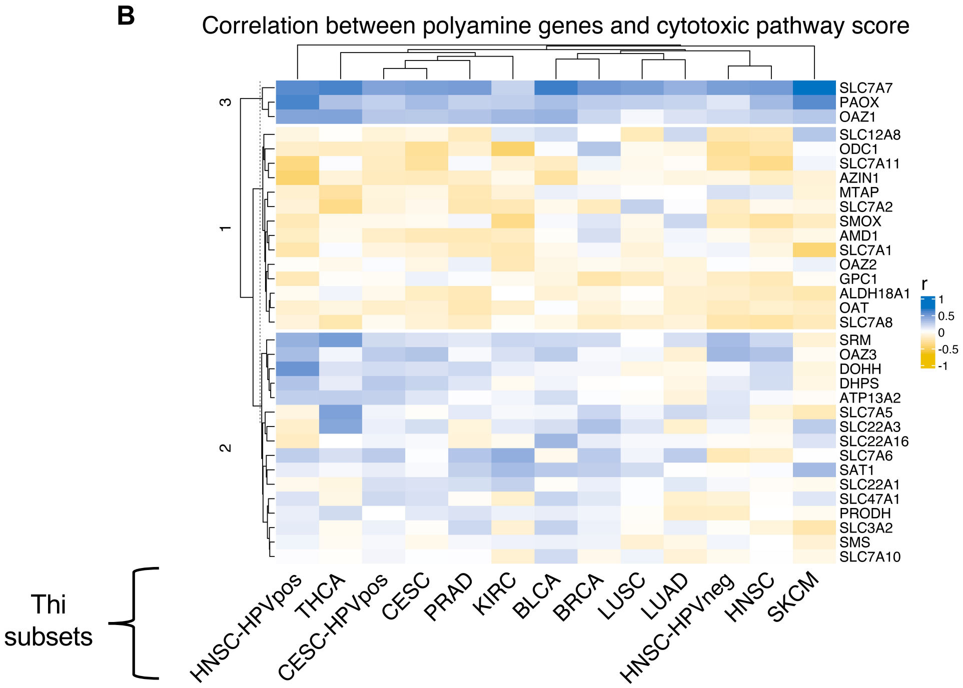


**Figure S3. Correlation between polyamine metabolism gene expression and ssGSEA scores for IFN*γ* and cytotoxic lymphocyte gene sets across TCGA Thi tumors.** (**A**) Results from pancancer analysis of correlation between polyamine gene expression and IFN*γ* gene set ssGSEA score. (**B**) Results from pancancer analysis of correlation between polyamine gene expression and cytotoxic lymphocyte ssGSEA score. *Blue,* positive correlation. *Gold*, negative correlation. Rows represent genes. Columns represent cancer type. Individual rectangles represent correlation coefficients for the correlation between gene expression and ssGSEA score across cancers.

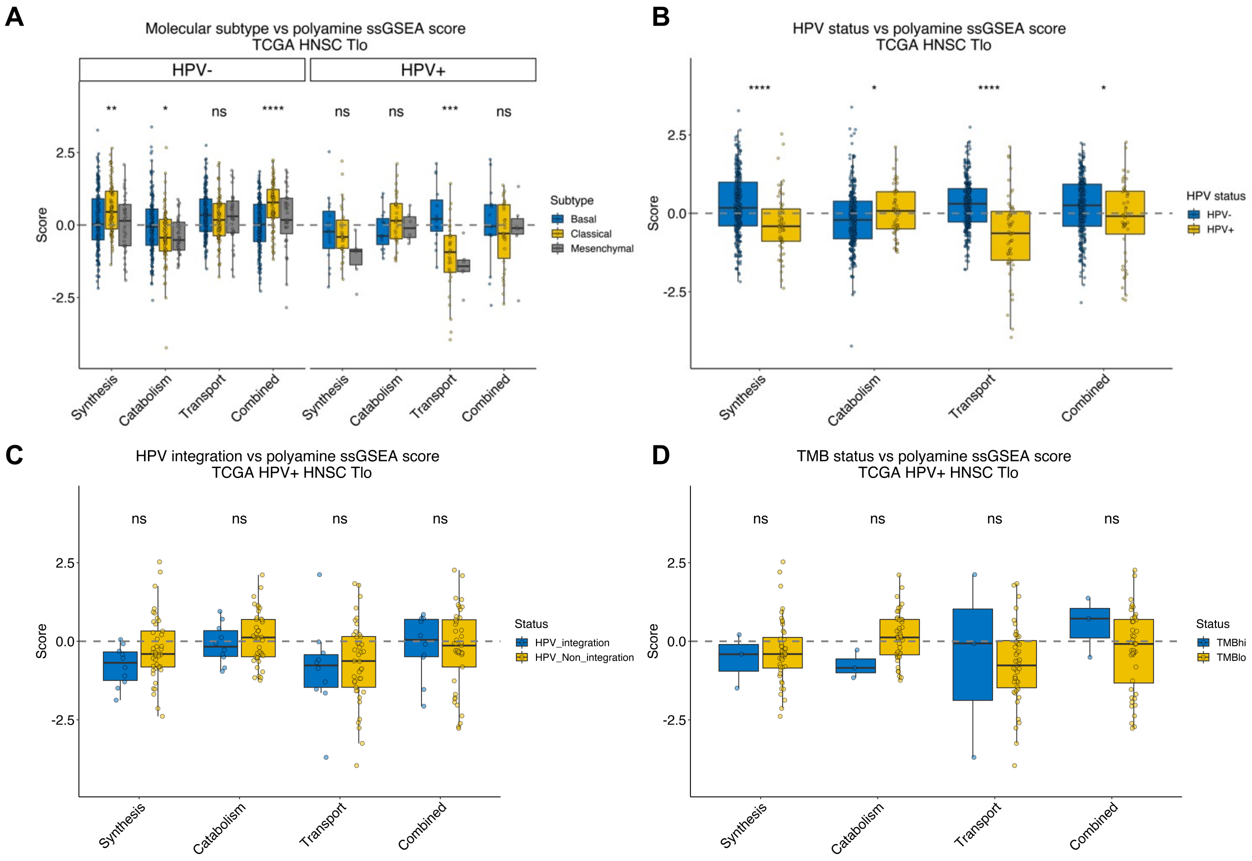


**Figure S4. Polyamine metabolism gene expression and tumor-intrinsic features among TCGA Tlo HNSCs.** Polyamine gene expression grouped by functional category and ssGSEA performed to generate scores. Polyamine gene set scores across TCGA Tlo HNSCs stratified by **(A)** molecular subtype, **(B)** HPV status, **(C)** HPV integration status as defined in Parfenov et al., and **(D)** tumor mutation burden (*TMBhi*, ≥ 10 mutations/Mbp; *TMBlo*, < 10 mutations/Mbp). *, p ≤ 0.05; **, p ≤ 0.01; ***, p ≤ 0.001; ****, p ≤ 0.0001; ns, p > 0.05.

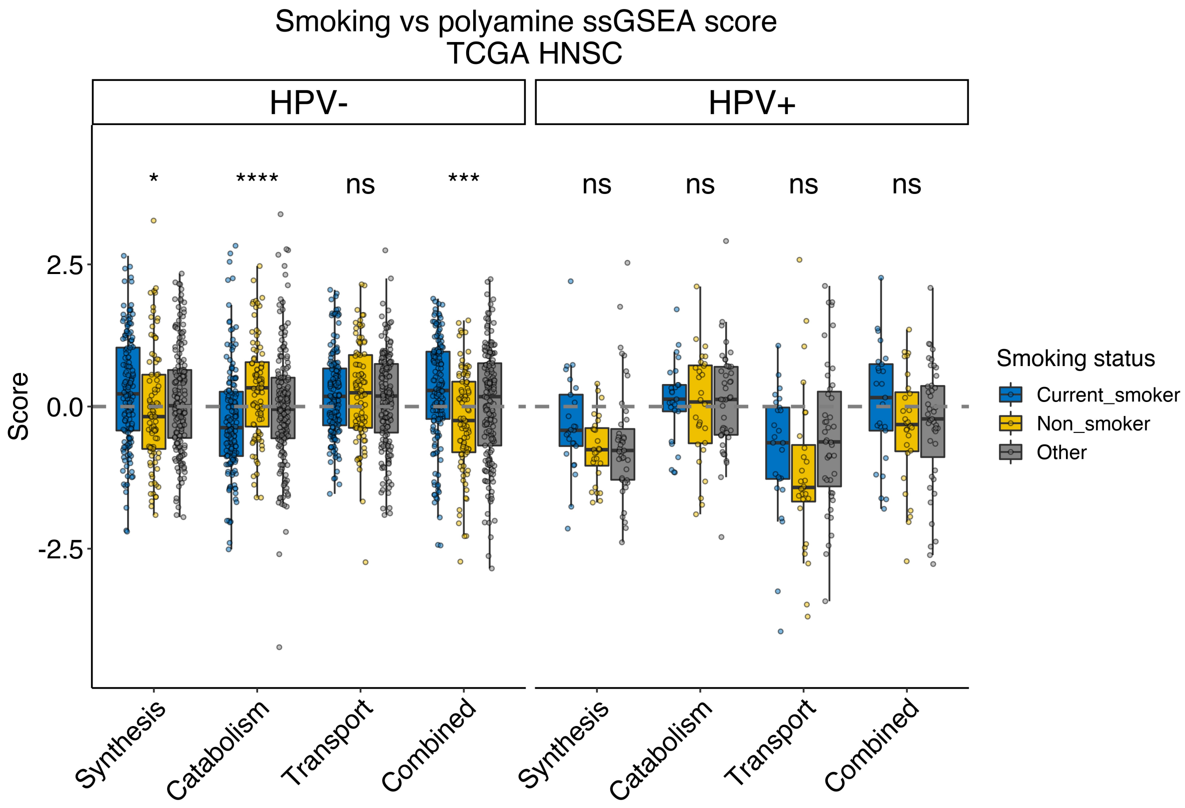


**Figure S5. Polyamine metabolism gene expression and smoking among TCGA HPV+ vs HPV- HNSCs.** Polyamine gene expression grouped by functional category. Polyamine ssGSEA scores are stratified by smoking status and plotted for HPV- and HPV+ HNSCs. *, $p \leq 0.05$; ***, $p \leq 0.001$; ****, $p \leq 0.0001$; ns, $p > 0.05$.

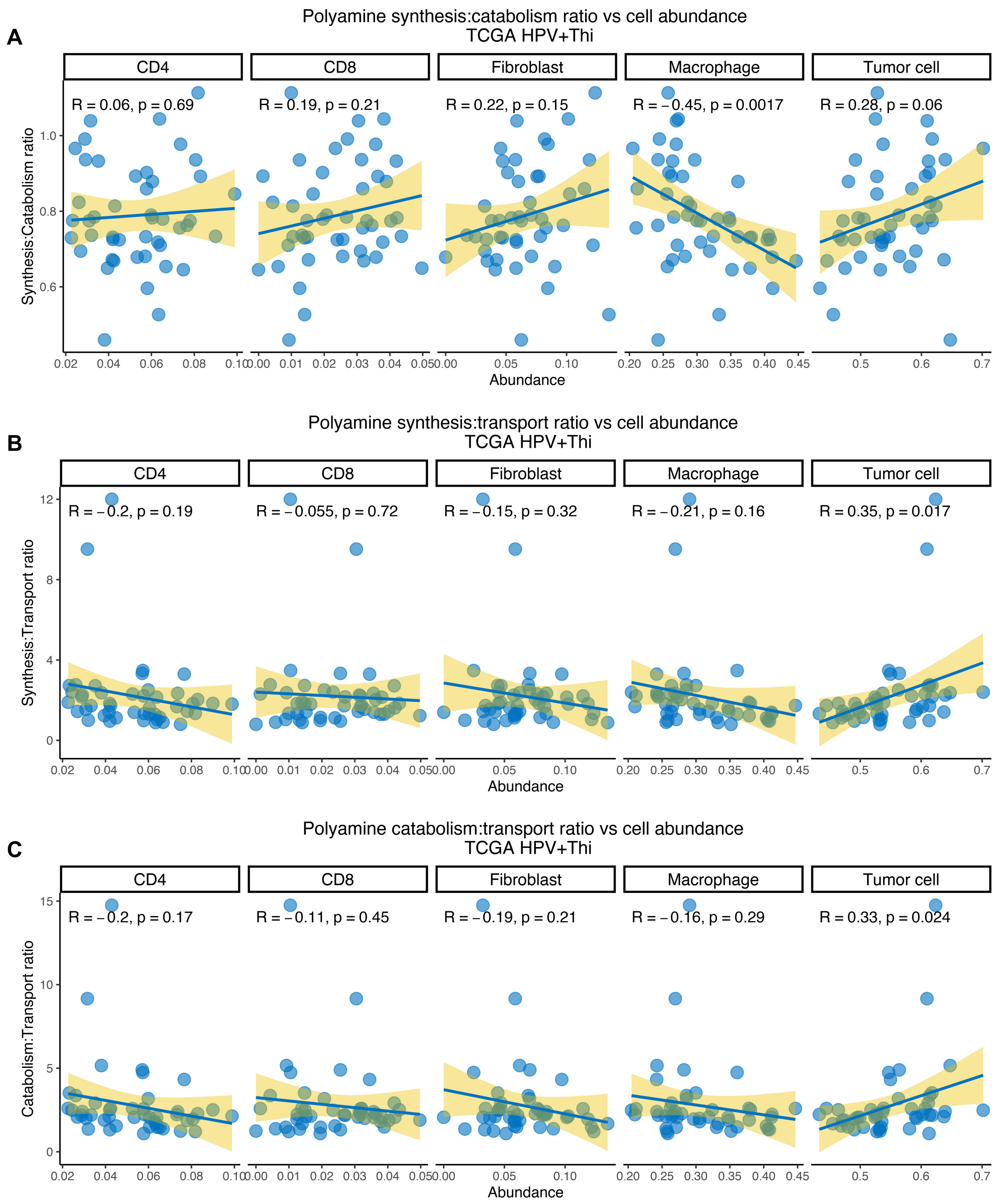


**Figure S6. Polyamine pathway expression ratios and cellular fractions in HPV+ Thi HNSC.** CIBERSORT-defined cellular abundance as a proportion of cell types in the tumor microenvironment. Cellular fractions stratified by polyamine **(A)** synthesis:catabolism, **(B)** synthesis:transport, and **(C)** catabolism:transport ratios. Relative cellular abundance is plotted on the x-axis and polyamine ssGSEA scores are plotted on the y-aixs. Pearson correlation coefficients and p-values are displayed in the plot windows.

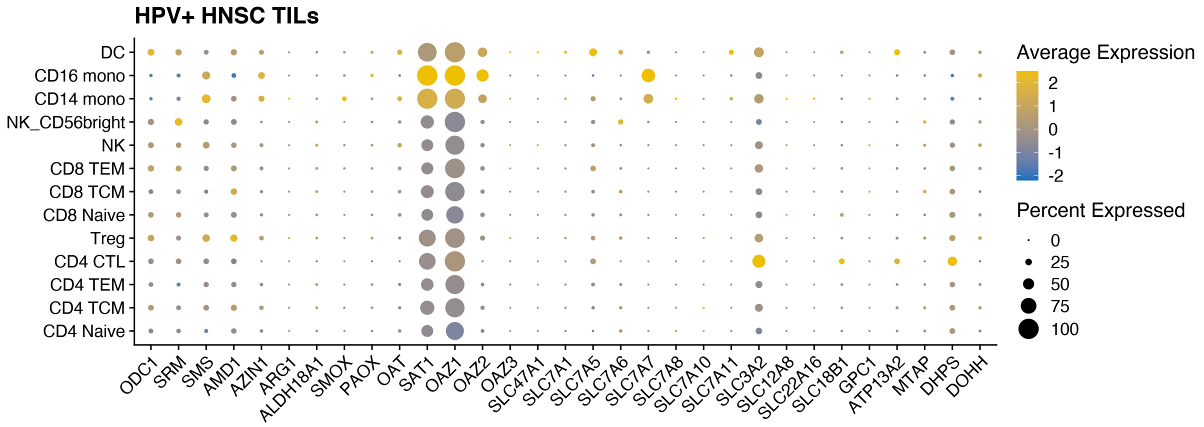


**Figure S7. Polyamine metabolism gene expression among tumor infiltrating lymphocytes in the HPV+ HNSC tumor microenvironment.** TIL single cell RNA sequencing data (Cillo et al.) from eight HPV+ HNSCs (14,859 cells) were mapped onto a reference single cell data set to infer cell lineage. Gene expression is plotted among inferred cell lineages. Scaled expression and percent of cells expressing each gene are represented on plot.

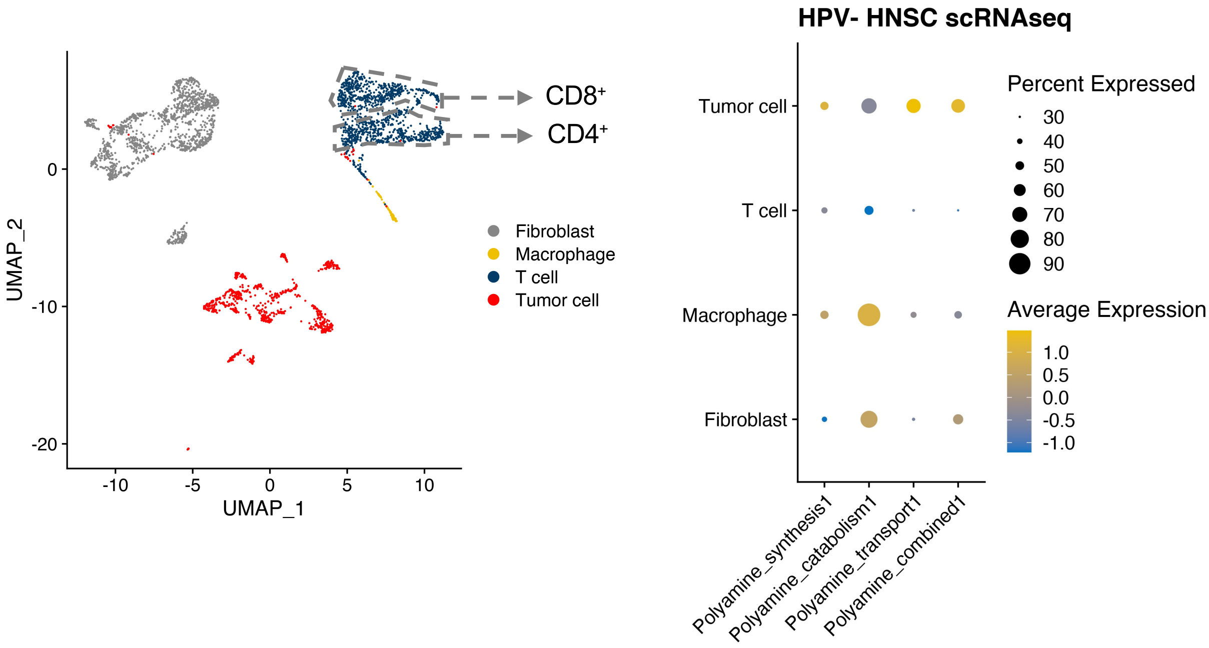


**Figure S8. Polyamine module expression in the HPV- HNSC tumor microenvironment.** (**A**) UMAP of single cell RNA sequencing data (Puram *et al*) from 21 HPV- HNSC tumor samples (3,478 cells) with cell lineage labeled according to the original classification. (**B**) Polyamine metabolism gene module expression was calculated and quantified by cell lineage.

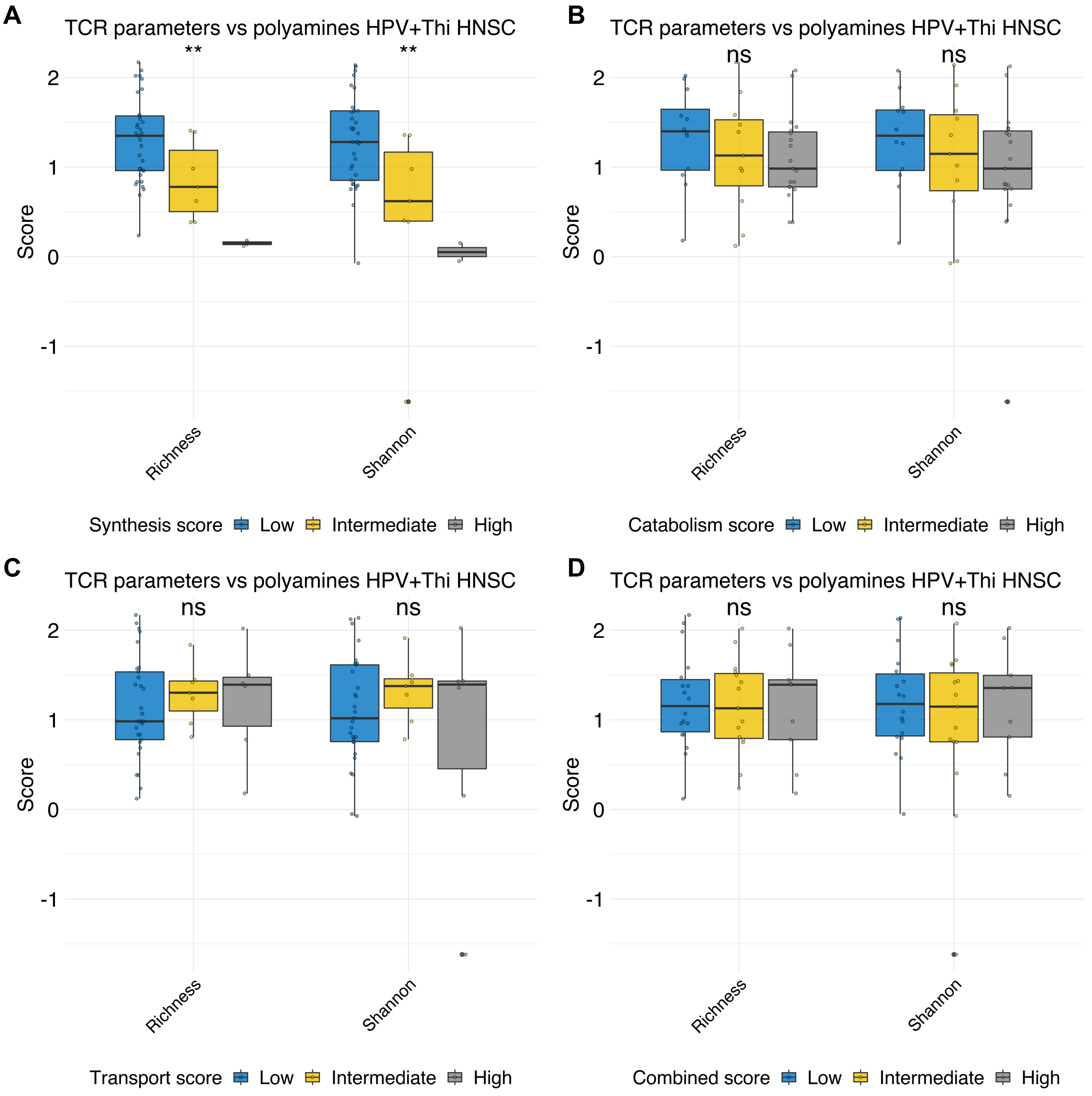


**Figure S9. Polyamine metabolism gene expression vs T cell receptor clonality richness and Shannon entropy among TCGA HPV+Thi HNSC.** Richness and Shannon scores from Thorsson et al. among TCGA HPV+Thi HNSCs (n = 97) were used. Stratified polyamine metabolism ssGSEA scores were used to assess for a relationship with TCR clonality. **, p ≤ 0.01.

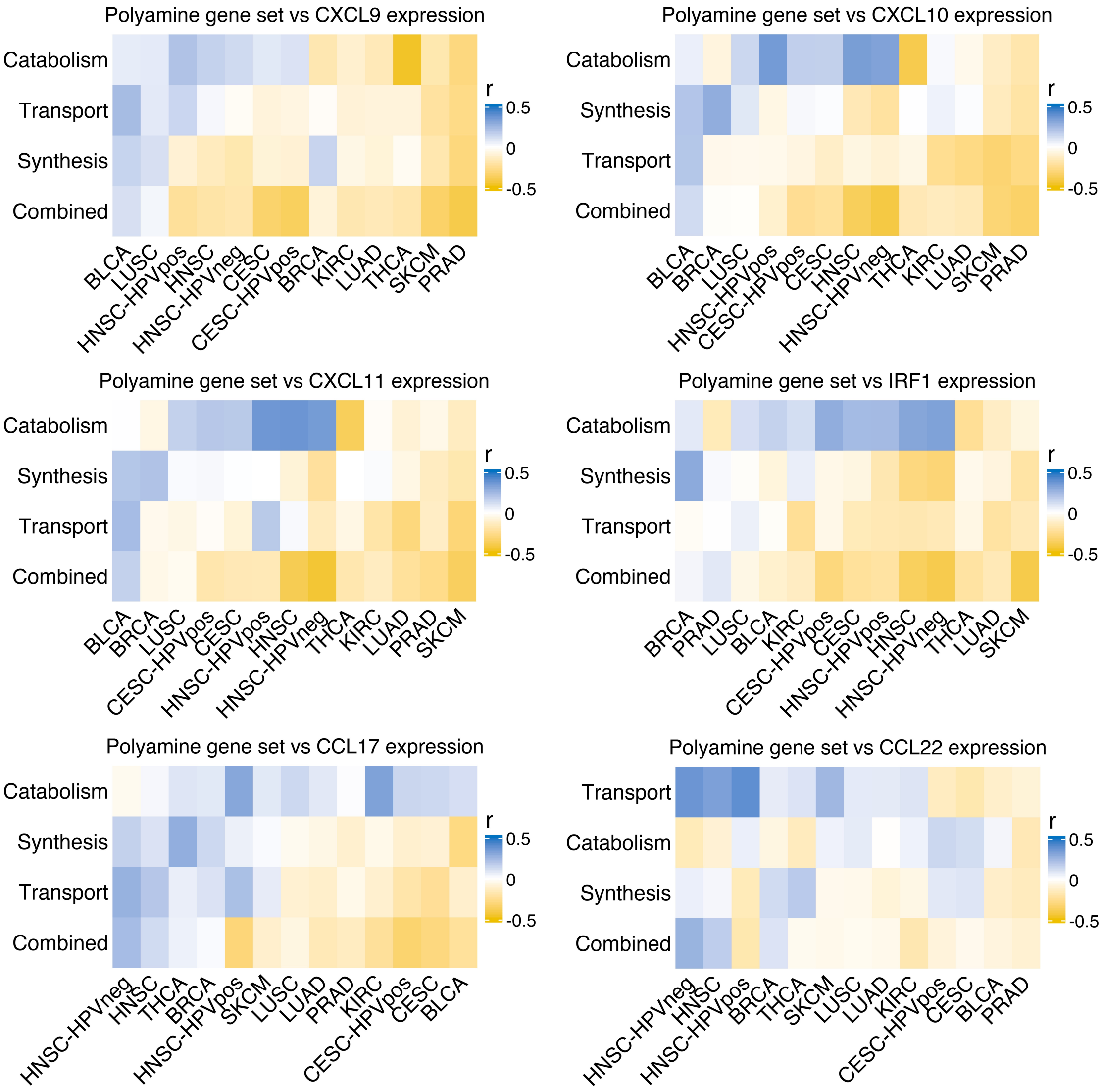


**Figure S10. Correlation between polyamine metabolism ssGSEA score and chemokines across cancers.** Correlation coefficients between polyamine ssGSEA scores and chemokine expression are shown in the heatmaps. Each rectangle represents a correlation coefficient for the correlation between polyamine ssGSEA score and chemokine expression for each cancer type.

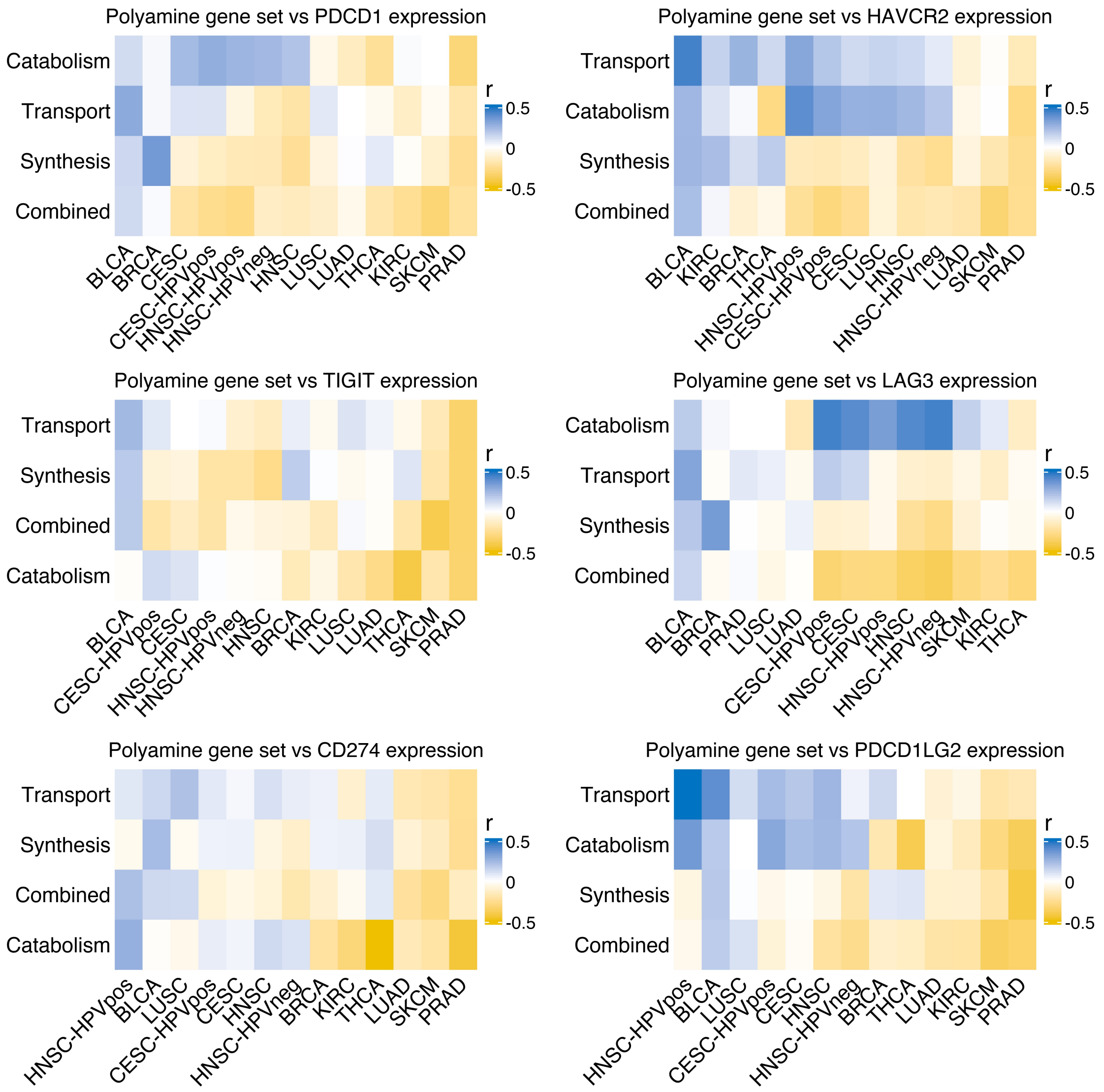


**Figure S11. Correlation between polyamine metabolism ssGSEA score and immune checkpoint genes across cancers.** Correlation coefficients between polyamine ssGSEA scores and immune checkpoint gene expression are shown in the heatmaps. Each rectangle represents a correlation coefficient for the correlation between polyamine ssGSEA score and immune checkpoint expression for each cancer type.

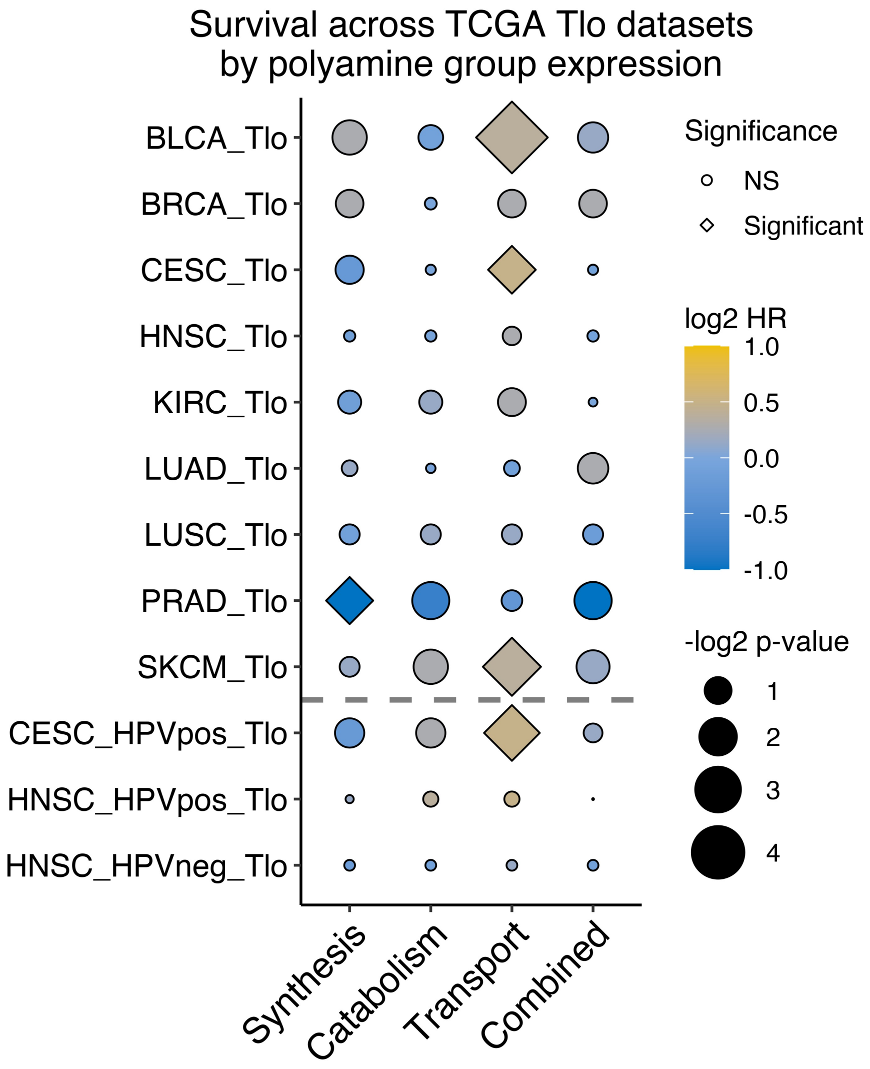


**Figure S12. Survival versus polyamine pathway gene set scores across T cell-depleted cancers.** Pan-cancer Cox proportional hazards analysis among the Tlo tumors for each cancer type was performed using dichotomized polyamine ssGSEA scores as a covariate. Log2 hazard ratios (HR) are plotted. Diamonds represent statistically significant associations (*Log test -log2 p-value,* FDR q < 0.25) and circles represent non-significant associations; size of shape represents magnitude of the q-value.
